# A Self-Explainable Dynamic Risk Monitoring Framework for Predicting Alzheimer’s Disease and Related Dementias

**DOI:** 10.1101/2025.10.20.25338232

**Authors:** Xiaoyang Ruan, Shuyu Lu, Sunyang Fu, Jaerong Ahn, Fang Chen, Rui Li, Andrew Wen, Liwei Wang, Ezenwa Onyema, Victoria Tang, Hongfang Liu

## Abstract

**Background:** Alzheimer’s Disease and Related Dementias (ADRD) affect millions worldwide and can begin over a decade before symptoms appear. ADRD are generally irreversible once clinical symptoms appear, making early prediction and intervention critical. While neuroimaging improves prediction, its availability restricts use at the population level. Electronic Health Record (EHR) data offers a scalable alternative, but existing models often overlook three key challenges: irregular clinical encounters, severe data sparsity, and the need for interpretability. To address these gaps, we propose GRU-D-RETAIN, a temporal deep learning architecture combines GRU-D’s strength in parameterized missing imputation with RETAIN’s explainable attention mechanism, enabling real-time risk monitoring at arbitrary clinical encounters with meaningful interpretations.

**Methods:** We identified 15,172 ADRD cases (age>=50) and 145,443 gender and date of birth matched controls from 6M patients in the University of Texas (UT) Physician EHR system. EHR were retrieved for each individual up to 10 years before ADRD diagnosis, and a random follow-up initiation date was assigned to simulate a real-world 10-year follow-up practice. Competing models including GRU-D-RETAIN, GRU-D, LSTM, Logit static, and Logit dynamic were trained on 6-fold cross-validation chunks and applied to the held-out to estimate performance.

**Results:** The scarcity of EHR records beyond 10 years before ADRD diagnosis precludes the development of valid predictive models beyond this timeframe. At the 10- year mark, only diagnoses of hypertension and hyperlipidemia exceeded 1% among ADRD cases. After randoming follow-up initiation date, GRU-D-RETAIN exhibited performance closely matching that of GRU-D across the entire follow-up period, both showing improved accuracy as follow-up time increases. Without applying data availability cut-off, both models achieved AUROC of 0.6 and 0.7 at 2-year and 8-year follow-up, respectively, significantly outperforming competing models. Data availability plays a more critical role than follow-up length in determining prediction performance. For example, 1 year of follow-up with 15% data availability yields comparable performance (AUROC of 0.75 and average precision of 0.5) to 7.5 years of follow-up with 10% data availability. For individual ADRD cases, GRU-D-RETAIN offered overall consistent explanations across training folds. However, certain folds produced different explanations at both the timestep and feature levels, despite yielding similar risk predictions.

**Conclusion:** We demonstrate that EHR data can support dynamic ADRD risk monitoring up to 10 years before diagnosis, though model utility depends highly on data completeness. GRU-D-RETAIN enables real-time risk monitoring with explainable attention weights at both timestep and feature levels, aiding clinicians in interpreting the output and identifying high-risk patients as well as potential key risk factors at individual level. This framework is broadly applicable to other conditions expecting irregular clinical encounters and requiring dynamic and interpretable risk assessment.

## Background

Alzheimer’s Disease Related Dementias (ADRD) impact tens of millions globally ^1^, an estimated 6.9 millions Americans age 65 and older today, and is the fifth leading cause of death among this age group ^2^. The brain changes associated with Alzheimer’s—such as amyloid-β accumulation and tau protein tangles—can begin more than a decade before noticeable dementia symptoms appear ^3^. During this early phase, individuals may remain cognitively normal but have increased risk of dementia progression. Currently all forms of AD/ADRD are considered incurable, and advanced stages often lead to life-threatening complications ^4^. Early detection of AD/ADRD offers an opportunity for timely therapeutic intervention, which may help slow disease progression and guide personalized care planning.

To date, numerous studies have been conducted on AD/ADRD prediction, either using Electronic Health Record (EHR) data alone ^5^, neuroimaging data alone ^6^, or both ^7^. While features from neuroimaging data greatly improve model performance ^6^, their limited availability ^8^, especially at the asymptomatic stage of ADRD, largely limited their application in the general population. As a comparison, routinely collected health data in EHR and clinical notes in the general population is a promising data repertoire. On this perspective, a large body of work has been revolving around EHR-based ADRD prediction ^5^, with various machine learning or deep learning models that take either static or temporal features to model various endpoints like probability of ADRD at a fixed future horizon ^9^ or next visit ^10^. However, almost none of the existing studies tapped simultaneously on two major challenges to the broader applicability of ADRD predictive models: 1) the extreme feature sparsity, particularly at asymptomatic stage of the disease, and 2) an elegant ante-hoc explainable model framework that can attribute the predicted ADRD risk to specific feature(s) at individual level to aid clinical interpretation.

On the data missingness side, existing studies primarily resorted to mean/median/mode imputation ^10,11^, semi complete-cases-only study ^9^, or not mentioned at all^12,13,14,15^. While these methods are easy to implement, they break the integrity of the model, wipe out informative missing ^16^ and introduce unmeasurable bias. This raises particular concern when asynchronicity and associated high volume of missing data are prevalent in the prolonged asymptomatic stage of ADRD. In terms of model explainability, most existing works did not provide direct model explanation ^10,17^, or resorted to post-hoc methods like SHAPLEY ^18^ or SHAP value ^9,14^, which rely on the assumption of feature independence that does not hold in temporally correlated data. It is also mathematically challenging and computationally prohibitive to calculate SHAP value on every time step for high dimensional longitudinal EHR with massive missingness. Moreover, research shows post-hoc explanation algorithms can produce inconsistent explanations for the same individual with the same model and dataset ^19^.

To this end, we propose a novel deep learning architecture named GRU-D-RETAIN to tackle the challenges associated with dynamic ADRD risk assessment. The proposed architecture combines the advantage of gated recurrent unit with decay (GRU-D) ^20^, which has proved robustness in automated missing parameterization and real-time risk monitoring by incorporating new measurements ^21,22,23^, and explainable reverse time attention architecture (RETAIN) ^24^, which can potentially provide feature and timestep level explanation. By incorporating advantages from both models, we prove that GRU-D-RETAIN can handle input features with varied data quality while providing reasonable explanations that align well with clinical knowledge.

## Materials and Methods

### Ethics Approval

The UT Institutional Review Board (IRB) approved the study (IRB Number: HSC-SBMI-24-0403), and the Ethics Committee waived the need for written informed consent from participants. The research was conducted in accordance with the Declaration of Helsinki and relevant institutional guidelines. This study involved secondary analysis of de-identified data and posed minimal risk to participants (Clinical trial number: not applicable).

### EHR databases

The study is based on the University of Texas (UT) Physicians EHR system, which includes a total of ∼6M patients as of 2025/01/01. The system was served by Allscripts before 2020, and then switched to EPIC. All records were mapped to the (Observational Medical Outcomes Partnership) OMOP common data model ^25^ before further analysis.

### Definition of ADRD diagnosis

In this study, we define ADRD diagnosis as the presence of concept code of ADRD or any other neurodegenerative cognitive diseases (Supplementary Table 1) along with either (1) use two or more records of dementia medication (Supplementary Table 2), or (2) at least one dementia medication record within 365 days of the diagnosis ^26^. Specifically for the second scenario, only the very first recorded dementia-related diagnosis in each patient’s history was considered. So patients with a standalone dementia-related diagnosis unaccompanied by any dementia-related medication within 365 days were considered ambiguous and excluded from analysis.

### Study cohort

#### Case group

From the 6M patients, we identified 81,985 patients with dementia-related diagnoses between 2000/01/01 and 2024/12/31. Among which 15,172 qualified our definition of ADRD diagnosis and 14,937 have age >=50 at index date were included into the case group (Figure 1).

**Figure 1:**
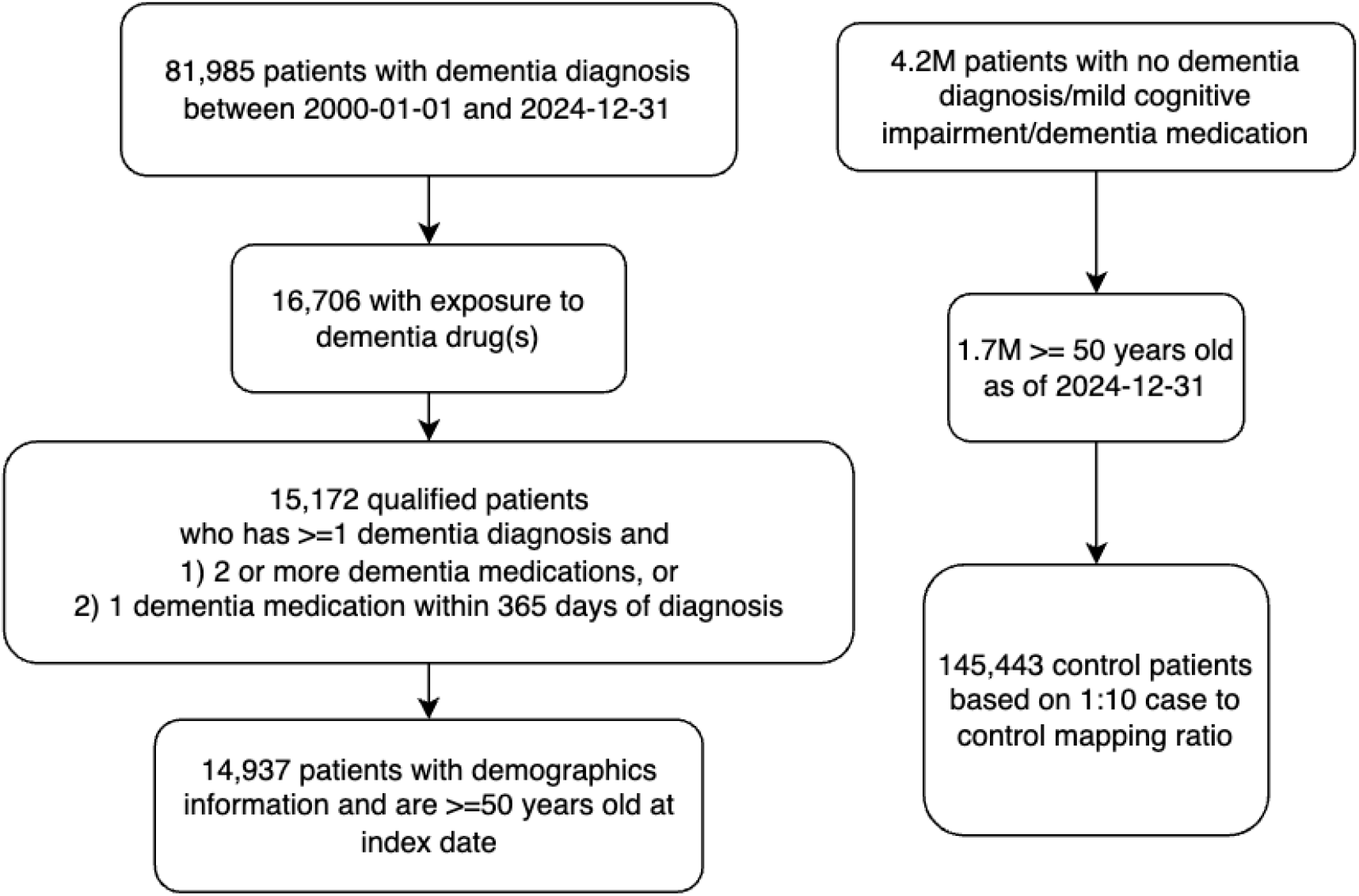
Construction flow for the ADRD case and control cohorts.

#### Control group

The control group was constructed from ∼4.2M patients that have no dementia-related diagnosis, no mild cognitive impairment, and no exposure to dementia-related medication. For each ADRD case, 10 control patients were matched on gender, race, ethnicity, and have a date of birth (DOB) within 30 days of the DOB of the corresponding case (Figure 1).

The demographics information of the case and control cohorts is shown in Table 1.

**Table 1:** Demographics of the ADRD case and matched control cohorts.

|  | ADRD Case | Control |
| --- | --- | --- |
| **Count** | 14937 | 145443 |
| **Age** |  |  |
| Median | 78 | 78 |
| Min | 50 | 50 |
| 1st Quartile | 72 | 71 |
| 3rd Quartile | 84 | 84 |
| Max | 106 | 107 |
| **Gender** |  |  |
| Female | 8293(56%) | 81517(56%) |
| Male | 5825(39%) | 57399(39%) |
| Unknown | 819(5%) | 6527(4%) |
| **Race** |  |  |
| White | 7951(53%) | 78630(54%) |
| Black | 2528(17%) | 24145(17%) |
| Asian <sup>a</sup> | 553(4%) | 4958(3%) |
| Others <sup>b</sup> | 28(0.2%) | 55(0.0%) |
| Unknown | 3877(26%) | 37655(26%) |
| **Ethnicity** |  |  |
| Non Hispanic | 9427(63%) | 92732(64%) |
| Hispanic | 1294(9%) | 11852(8%) |
| Unknown | 4216(28%) | 40859(28%) |

<sup>a</sup> Includes Chinese, Japanese, Korean, Filipino, Vietnamese, Asian Indian
<sup>b</sup> Includes American Indian or Alaska native, Other pacific islander, and Polynesian

### Definition of index date and endpoint

The index date for ADRD diagnosis was defined as the earliest occurrence of either a dementia-related diagnosis or the first exposure to a dementia medication, whichever first. For control subjects, the index date was aligned with the index date of their matched case. In the remaining text, we refer to an endpoint (or end of follow-up) as when an ADRD case or control reached his/her index date.

### Definition of follow-up initiation date

In real-world clinical settings, the exact date of ADRD diagnosis (i.e., the index date) is unknown in advance. Therefore, using “10 years before the index date” as the start of follow-up is not feasible. To better reflect real-world conditions—where patients may begin clinical follow-up at a random time point—we assigned each individual in both the case and control groups a randomly selected follow-up initiation date. The model is only allowed access to data from the start of follow-up until just before the index date; any data prior to the follow-up initiation or after the index date remains hidden (see Figure 2).

**Figure 2:**
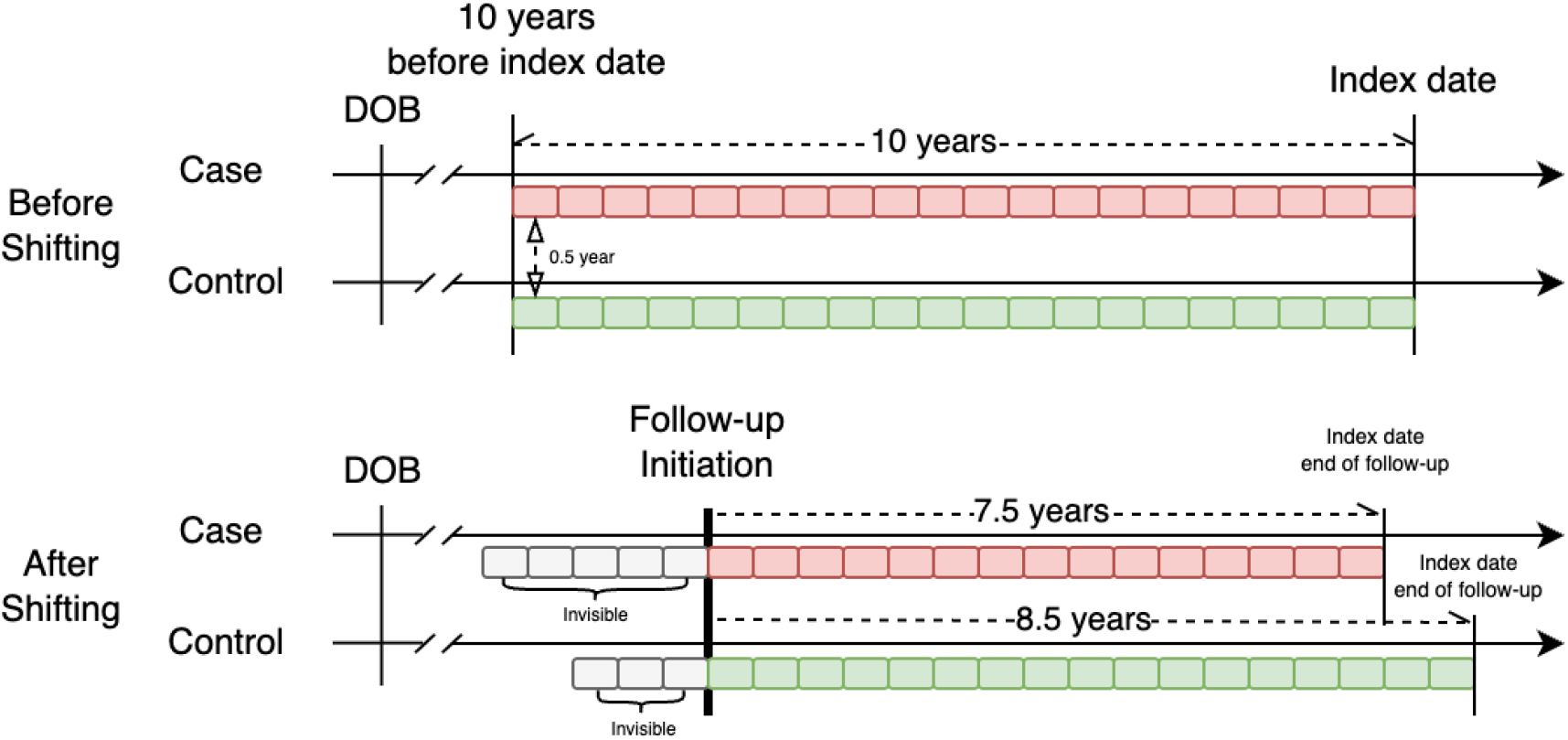
Illustration of random shifting and follow-up initiation date. Red and green tiles indicate data from cases and controls, respectively, during the 10-year period leading up to the index date. Following the shift, grey tiles represent temporal data hidden from the model during both training and prediction. In this example, the ADRD case and control have 7.5 and 8.5 years of follow-up, respectively, prior to reaching the index date.

### Definition of outcome

Outcome is defined as the probability of ADRD diagnosis within 10 years from follow-up initiation.

### Input features and data preprocessing

#### Diagnosis code

A total of 4,645 unique ICD-9 diagnosis codes were grouped into 265 Clinical Classifications Software (CCS) categories, as defined by the Healthcare Cost and Utilization Project (HCUP) ^27^. All ICD-10 diagnosis codes were mapped to ICD-9 codes using the General Equivalence Mapping provided by the Centers for Medicare & Medicaid Services. CCS provides a framework for aggregating ICD-9 diagnoses and procedures into a smaller number of clinically meaningful categories to facilitate interpretation and statistical analysis. The resulting CCS codes were one-hot encoded for input into the machine learning models.

#### Lab and Vital measurements

Laboratory and vital sign measurements were extracted from the OMOP measurement table, covering up to 10 years prior to the index date for each patient. To reduce input feature sparsity, we selected only those measurements with a yearly presence rate greater than 5% in the case group during any of the 10 years. Measurement values were standardized using z-score normalization, with the mean and standard deviation calculated from the training set. These parameters were then applied to the validation, testing, and held-out datasets. Normalized z-scores were clipped to a range between -5 and 5 before being input to the machine learning models.

#### Demographics

Static demographic variables—such as gender, race, and ethnicity—were one-hot encoded and replicated across all time intervals. Age was treated as a dynamic variable that increases over time and was normalized by dividing by 100.

#### Procedures

Procedure records were extracted from the OMOP procedure_occurrence table, covering up to 10 years prior to the index date. Concept codes were mapped to seven major SNOMED procedure categories: abdomen, cardiac, intracranial, joint, skin, spine, and thoracic (Supplementary Table 3). For each patient, the total procedure duration (in hours) within each time interval was calculated separately for each of the seven categories and used as input features.

#### Risk medications

Risk medications refer to drugs that have been reported to potentially increase the risk of developing dementia. We included 187 such medications, categorized into 12 major classes, including anticholinergics, anticonvulsants, antidepressants, among others (Supplementary Table 4). Only records with 30-days or longer exposure length were considered. Exposure to these medications was encoded for machine learning models as binary values—1 indicating exposure and 0 indicating no exposure—across defined time intervals.

#### Functional Status Ascertainment (FSA)

FSA was conducted with FedFSA, a hybrid federated AI framework designed to extract functional status information from longitudinal EHRs. It identifies impairment using six categories for basic activities of daily living (ADL) and nine for instrumental ADLs (IADL), based on the International Classification of Functioning, Disability and Health (ICF) standard ^28,29,30^. The framework includes open-source components: an information extraction pipeline built on Apache Unstructured Information Management Architecture (UIMA), a federated learning engine, and a large language model. Its design separates knowledge sources (e.g., keywords, rules) from NLP processes, improving portability. FedFSA is compatible with the Open Health NLP (OHNLP) Backbone ^31^, allowing scalable Extract-Transform-Load (ETL) workflows for easier adoption. Local evaluation on 68 patients from a geriatric osteoporosis clinic showed strong performance across 15 ADL/IADL categories: precision 0.868, sensitivity 0.828, specificity 0.937, NPV 0.949, and F1 score 0.816.

#### Sampling scheme of dynamic variables

Based on a systematic review of the time intervals between measurements and considering a clinically practical frequency for ADRD risk assessment, we selected a six-month interval for sampling dynamic variables (i.e., variables that change over time). This resulted in 21 time steps covering the 10-year period prior to the index date.

### Deep learning models GRU-D model

The basic architecture of the GRU-D model has been systematically described by ^20^, here we recapitulate the equations for handling missing values.

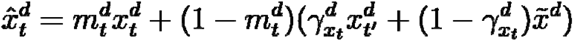

where 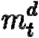 is the missing value indicator for feature *d* at timestep *t*. 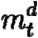 takes value 1 when 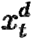 is observed, or 0 otherwise, in which case the function resorts to weighted sum of the last observed value 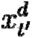 and empirical mean 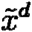 calculated from the training data for the *d* th feature. Furthermore, the weighting factor 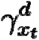 is determined by

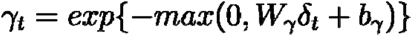

where *W*_*γ*_ is a trainable weights matrix and *δ*_*t*_ is the time interval from the last observation to the current timestep. When *δ*_*t*_ is large (i.e. the last observation is far away from current timestep), *γ*_*t*_ is small, results in smaller weights on the last observed value 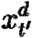, and higher weights on the empirical mean 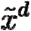 (i.e. decay to mean).

### GRU-D-RETAIN model

The GRU-D-RETAIN model architecture (Figure 3) is rooted on GRU-D, which provides automated missing imputation, and extended with architecture similar to the interpretable reverse time attention architecture named RETAIN as described by ^24^. In GRU-D-RETAIN, the actual observation or otherwise imputed value 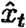 is multiplied with *β*_*t*_ and *α*_*t*_, the feature wise and time step wise partial coefficient at time step t to obtain context vector *c*_*t*_. *c*_*t*_ from multiple historical timesteps are then summed up and casted to output 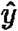 through output vector weights matrix 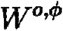. For each output dimension, the interpretability for a specific timestep t comes from *α*_*t*_*β*_*t*_*W*^.,*ϕ*^ as partial coefficient of 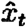.

**Figure 3:**
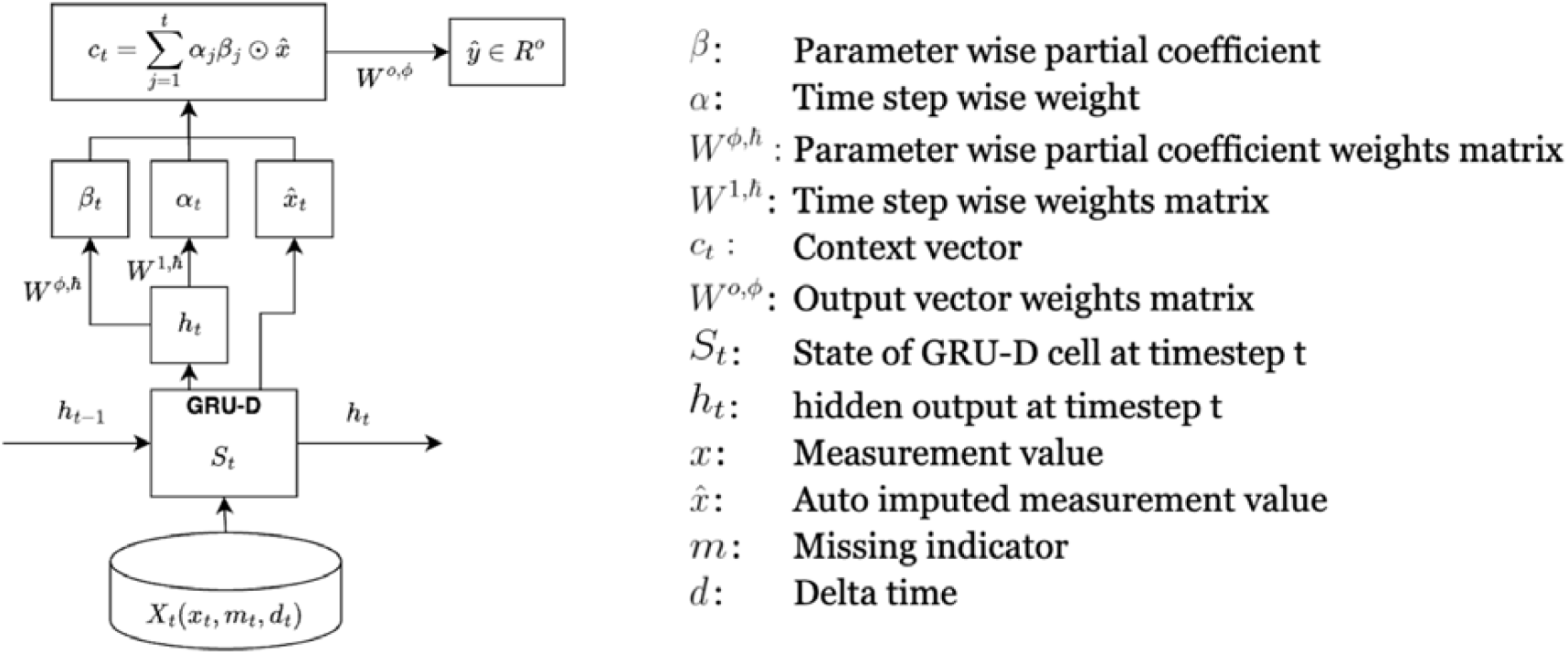
GRU-D-RETAIN architecture

### Hyperparameters and training schemes

The hyperparameters and training schemes were manually determined by varying combinations of hidden neuron size (10-200), learning rate (0.001-0.1), batch size (10-50), drop out ratio (0-0.5), optimizer (SGD, RMSprop, ADAM), regularization scheme(L1/L2), Beta coefficients smooth out threshold (1, 5, 10), momentum (0-0.5), and early stopping thresholds (1%-10% error differences between training and validation dataset) that optimize the performance of GRU-D-RETAIN on non held-out datasets.

### Long Short-Term Memory (LSTM) model

The LSTM model consists of a single LSTM layer with a hidden size of 128, followed by fully connected layers. Input features underwent the same preprocessing pipeline as the logistic regression models. Missing values were first handled using the Last Value Carried Forward (LVCF) method. For computational efficiency, any remaining missing values were then filled using simple mean imputation. The LSTM model was trained using the same six-fold cross-validation setup as the GRU-D-like models. The LSTM model processes the entire 21-timestep sequence and generates predictions at each step simultaneously. At each timestep, records that had already reached the endpoint were excluded for the subsequent timesteps.

### Logistic Regression

Logistic regression models with L1 regularization (Lasso) were trained using the same six training folds as the deep learning models and applied to the held-out fold at each of the 21 timesteps. At each timestep, records that had already reached the endpoint were excluded. For the remaining records, missing values were first handled using the LVCF method. Any remaining missing values were then imputed using an iterative imputation method with a decision tree regressor, allowing up to 20 iterations.

Two modeling strategies (Supplementary Figure 1) were evaluated at each timestep: Logit Dynamic and Logit Static. The Logit Dynamic approach trained 21 separate logistic regression models, each using the available data at the respective timestep after applying random shifting. In contrast, the Logit Static approach followed a conventional case-control design, training a single logistic regression model using data from ADRD cases and matched controls at the endpoint.

### Evaluation Metrics

We assessed model performance using established metrics, including the area under the receiver operating characteristic curve (AUROC), average precision (defined as the weighted average precision across all sensitivity levels), and the F-score (computed at the optimal threshold that maximizes the geometric mean of sensitivity and specificity). In addition, we incorporated two clinically relevant metrics: flag rate, defined as the proportion of patients flagged to achieve 60% sensitivity across all remaining ADRD cases within 10 years following follow-up initiation; and the percentage of cases identified in the next half year, defined as the proportion of true ADRD cases—among those predicted to be in the 3rd and 4th risk quartiles—that have ADRD diagnosis within the subsequent six months. All metrics were computed at each of the 21 timesteps, excluding patients who had already reached the endpoint prior to the timestep of interest.

### Permutation Feature/Modality Importance

Permutation feature importance was assessed at both the feature and modality levels using the held-out dataset. For each targeted feature or modality, sample IDs were randomly shuffled to disrupt the original associations. The AUROC was calculated at each of the 21 timesteps before and after permutation, and the average percentage change—computed as (before − after) / before × 100%—was used to quantify the model’s reliance on that feature or modality. One permutation was conducted per training fold, yielding six data points per feature/modality to support confidence interval estimation.

### Partial Dependence Plot

Partial dependence plot (PDP) was conducted to assess how predicted ADRD risk changes as input feature value was manually tuned, thus estimating the direction of impact for reference in clinical intervention. The tuning was conducted by incrementing or decrementing each feature of each of the 2,070 ADRD cases in the held-out dataset for the following modalities. Lab and vital measurements (increase/decrease 1 standard deviation after z-score normalization), CCS codes (present/absent), risk medications (present/absent). At each follow□up timestep, we recorded the changes in ADRD risk probabilities induced by these tunings (versus no tuning), then averaged the differences across all valid timesteps for each feature of each ADRD case. FSA, procedures, and demographics were excluded from this analysis due to either extremely low data availability or being considered not amenable to intervention.

### Computing Platform

Data analysis was conducted on a Unix system equipped with 96 CPU cores (Intel® Xeon® Gold 6442Y) and 1.0□TB of memory. Data curation and visualization were carried out using R version 4.3.1. Deep learning tasks were implemented in Python 3.12.3 with PyTorch 2.5.1. Lasso regression was performed using the Lasso module from scikit-learn 1.7.0, and missing data imputation for the Lasso models was handled using the DecisionTreeRegressor module from the same scikit-learn version.

## Results

### Study population

The study included 14,937 individuals with ADRD diagnosis and 145,443 age and gender matched controls. Participants ranged in age from 50 to 107 years old, with females comprising 56% of the ADRD case group. The ADRD and control groups had comparable distributions of age, gender, race, and ethnicity (Table 1).

### Data availability before ADRD diagnosis

Analysis of the five modalities revealed very limited data availability, measured as yearly feature presence rate, prior to 10 years before ADRD diagnosis. For instance, among the 756 types of vital signs and lab tests recorded in the EHR for current ADRD cases, the highest yearly presence rate before the 10-year mark was below 0.52% (i.e. only about one in 200 patients had a measurement recorded in a year). After applying a feature presence rate threshold, we retained 64 out of 756 vital and lab features, and 81 out of 281 CCS diagnosis codes, each with at least a 5% yearly presence rate, for use in model training and prediction. Along with the FSA score, risk medications, and procedures, the final list of features used by the models and their presence rate over time are visualized in Supplementary Figure 2 and tabulated in Supplementary Table 5. In general, the presence rates of all features increased over time, peaking at the time of ADRD diagnosis.

### Performance of GRU-D-RETAIN in dynamic ADRD risk assessment

To simulate real-world conditions in which patients may initiate ADRD risk assessment at varying time points, we randomly assigned a follow-up initiation date to each individual in the study cohort, and masked the data prior to the initiation date (Figure 2). The feature presence rate after random shift is visualized in Supplementary Figure 3. As illustrated in Figure 4, both GRU-D and GRU-D-RETAIN demonstrated comparable performance across the entire follow-up period and showed improved predictive accuracy on longer follow-up time. Notably, a minimum of 2 years of follow-up was needed to reach an AUROC of 0.6, whereas 8 years were required to achieve an AUROC of 0.7. Over the 10-year follow-up period, 60% to 75% of individuals with above-median predicted ADRD risk were diagnosed with ADRD within the subsequent six months. In contrast, the LSTM model generally underperformed GRU-D and GRU-D-RETAIN on major metrics throughout the 10-year follow-up. Atemporal logistic regression models were unable to demonstrate clinical utility, with AUROC values remaining near 0.5 and flag rates around 0.6 throughout the follow-up period.

**Figure 4:**
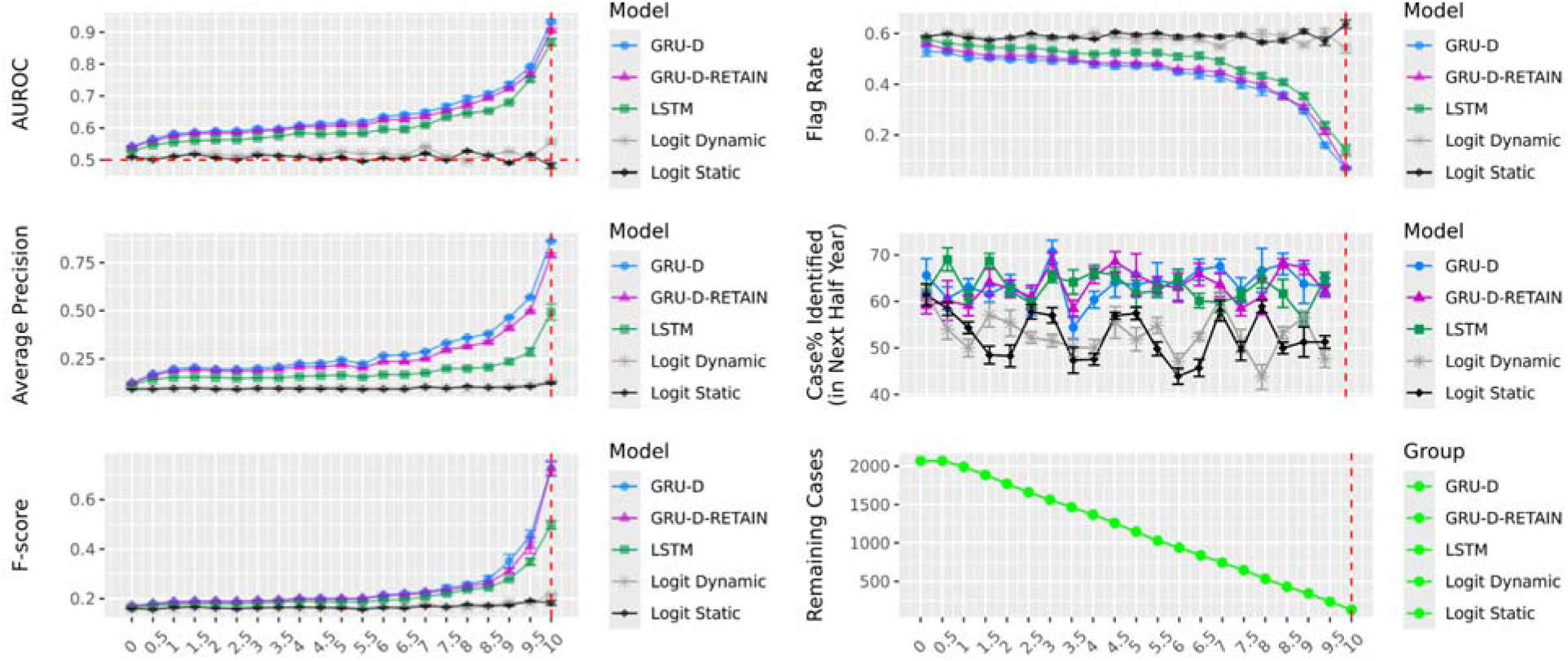
ADRD risk prediction performance throughout 10-year follow-up. Predictions made on the held-out dataset. Flag rate the lower the better. Error bars represent 95% CI based on 6-fold models.

### Impact of data availability on prediction performance

At various follow-up durations, greater data availability is associated with improved prediction performance (Figure 5). A significant performance gain was observed when data availability—measured by the average yearly feature presence rate within the 10 years preceding ADRD diagnosis—exceeds 10%. Notably, model performance can be enhanced either by extending the follow-up period or by increasing data availability, with the latter playing a major role. For instance, in terms of AUROC and average precision (i.e. the proportion of true positives among predicted positives), 1 year of follow-up with 15% data availability yields performance (AUROC of 0.75 and average precision of 0.5) comparable to 7.5 years of follow-up with 10% data availability.

**Figure 5:**
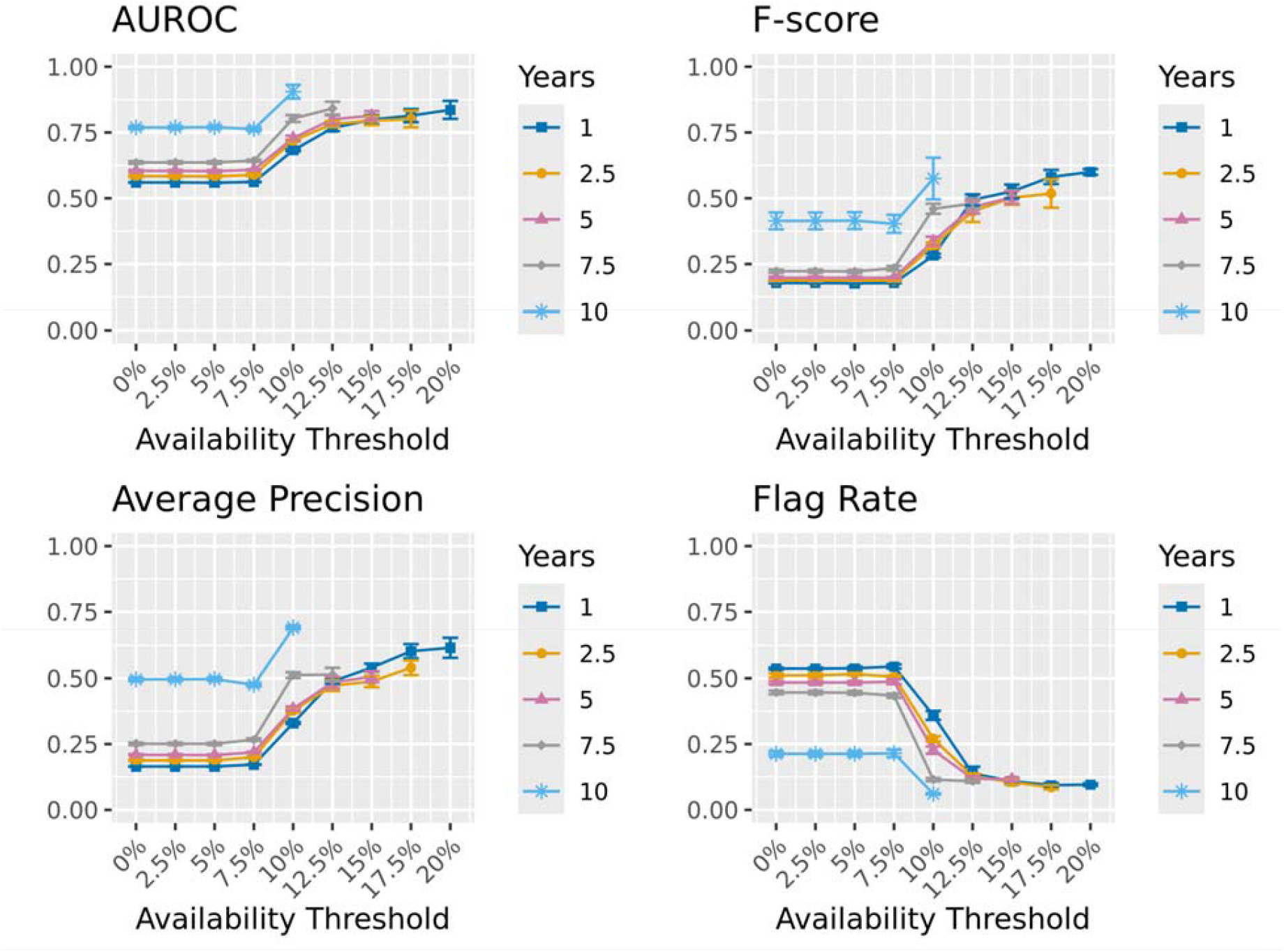
Prediction performance across varying data availability thresholds and follow-up durations. For each threshold, ADRD cases with data availability exceeding the specified level were included, along with their matched controls (regardless of the controls’ data availability). Metrics calculated for a minimum of 30 ADRD cases. Error bars represent 95% CI based on 6-fold models.

### Permutation Modality and Feature Importance

At the modality level, CCS codes showed the greatest impact, with permutation leading to an average AUROC decrease of 12.4% (95% CI: [11.7%–13.2%]). The remaining modalities, ranked by decreasing impact, were demographics (2.8%; [2.3%–3.4%]), lab and vital measurements (1.0%; [0.8%–1.2%]), risk medications (0.9%; [0.5%–1.3%]), FSA (0.2%; [0.1%–0.3%]), and procedure length (0.03%; [–0.003%–0.1%]).

Figure 6 presents the top 20 features that led to the greatest AUROC reduction following permutation. Among the 81 CCS codes entered in the model, the most influential one was the diagnosis of delirium, dementia, and other cognitive disorders, which caused a 3.1% (95% CI: [2.7%–3.5%]) decrease in AUROC. The second important CCS code was unclassified residual codes (0.8%; [0.3%-1.2%]). Significant or marginally significant effects were also observed for diagnoses such as other nervous system disorders, hyperlipidemia ^32^, mood disorders ^33^, Parkinson’s disease ^34^, spondylosis/intervertebral disc disorders/other back problems ^35^, osteoarthritis ^36^, and hypertension ^37^ among other CCS codes (Supplementary Figure 4). Most demographic factors showed significant influence, with gender, race, and ethnicity showing more contribution than age (Figure 6). The use of antipsychotic ^38^, anticonvulsant/antiepileptic ^39,40^, and antihistamine ^41^ medications showed significant effect (Supplementary Figure 5). In contrast, antidepressant ^42^ and anticholinergic ^43^ only have marginal impacts. Labs and vital measurements generally have <0.1% impact, with factors like alanine aminotransferase ^44 45^, mean corpuscular volume ^46 47^, respiratory rate ^48^, thyrotropin ^49^, and urea nitrogen ^50^ showed significance (Supplementary Figure 6). For FSA scores, only difficulty in self managing medication showed significance (Supplementary Figure 7). Among the seven procedure types, joint procedure ^51^ has the largest effect size (<0.05%) but failed to show significance. Notably, no features were found to improve AUROC after permutation, suggesting an absence of confounding or misleading signals in the model.

**Figure 6:**
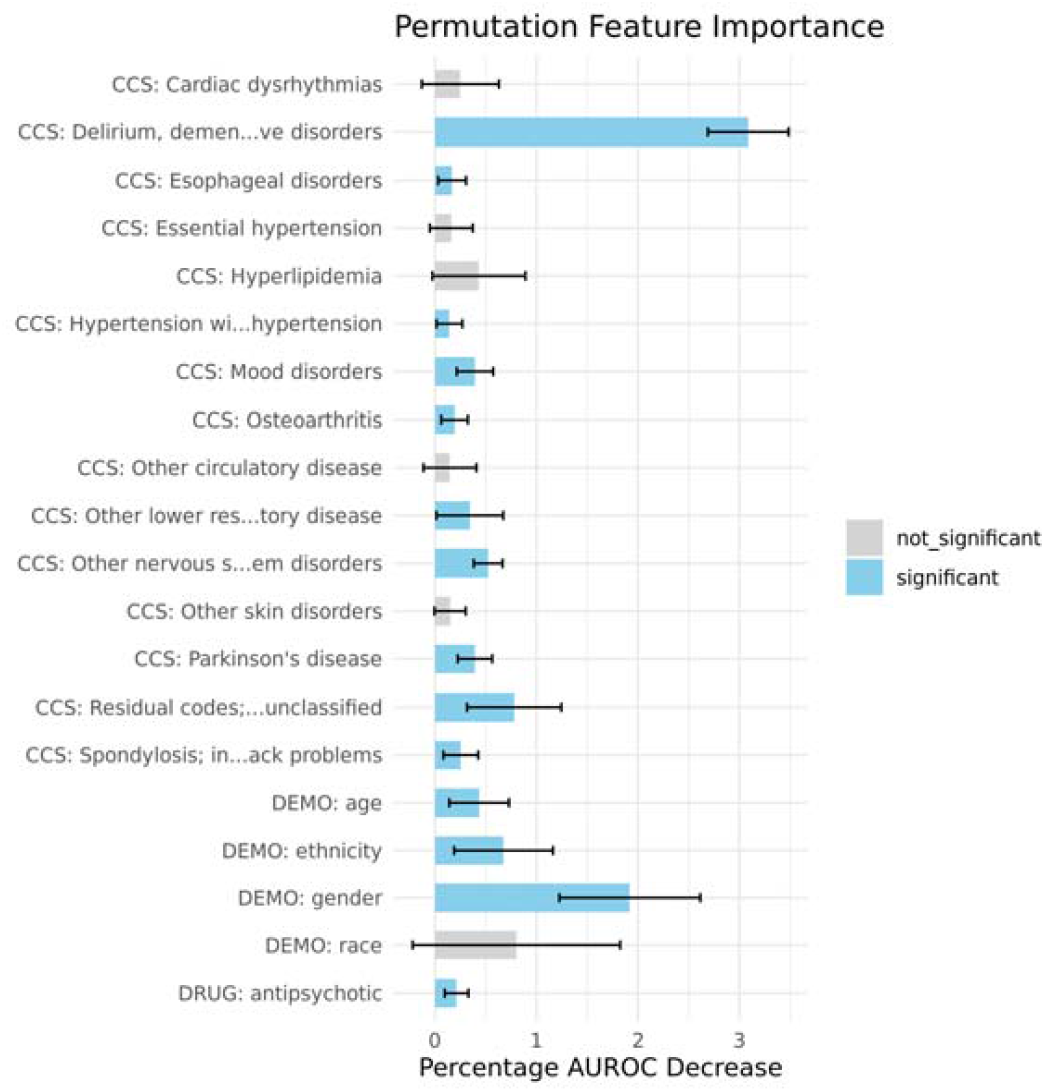
Top 20 features contributing to AUROC. Error bars represent 95% CI based on 6-fold models.

**Figure 7:**
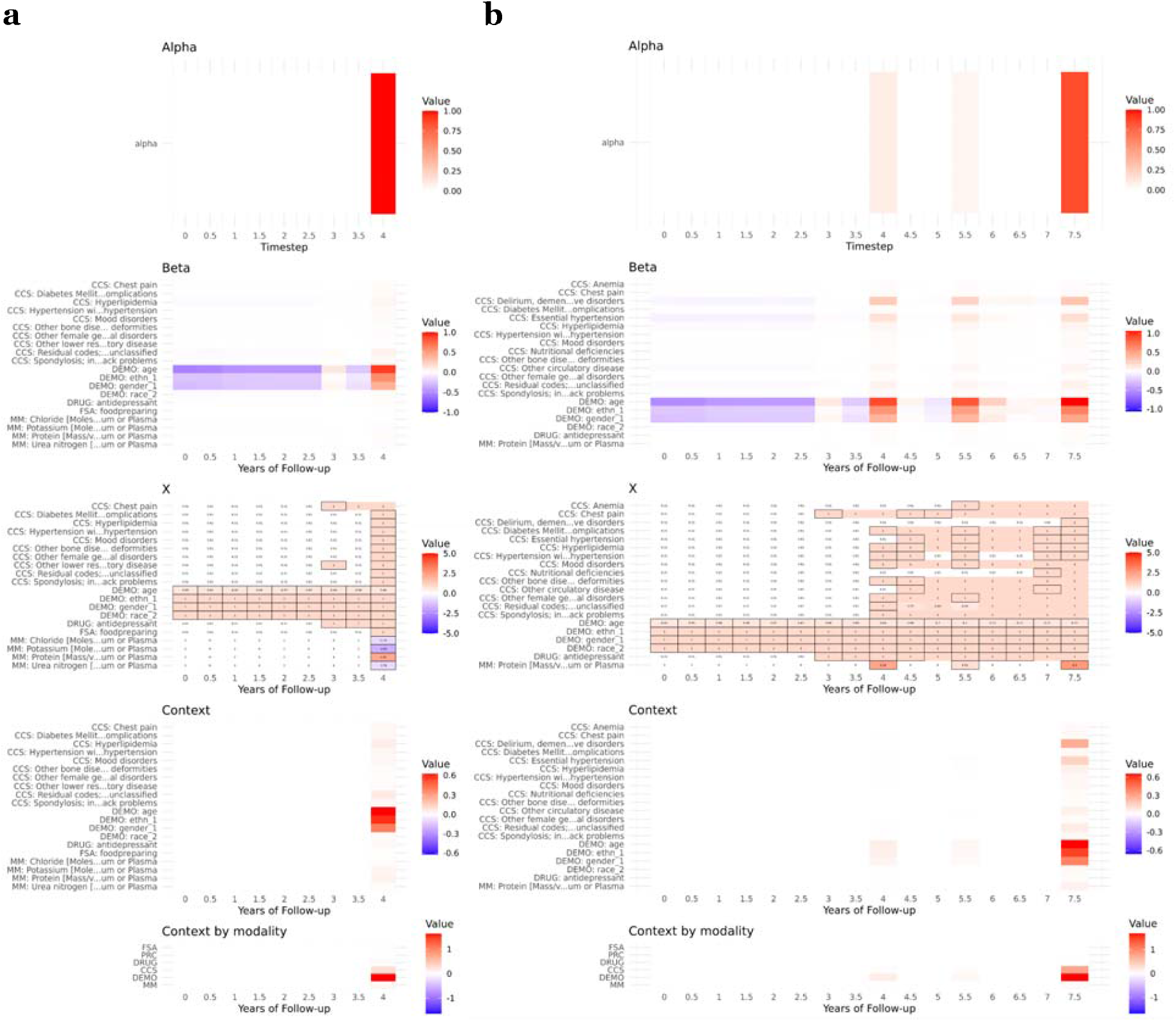
Interpretation for patient A with ADRD diagnosis at the 7.5th year of follow-up. a) Interpretation at 4 years, where the predicted risk significantly increases above the previous time interval. b) Interpretation at 7.5 years, where the patient had an ADRD diagnosis. Alpha: timestep level attention; Beta: feature level attention; X: values of input features; Context: Alpha*Beta*X. For input features (X), cells in black border indicate actual observed values, while others indicate missing observations imputed by GRU-D on the fly. Data shown for the top 20 features with largest contribution to the predicted risk at corresponding timestep.

### Partial dependence plot

Partial dependence plots for laboratory and vital measurements, CCS codes, and risk medications are shown in Supplementary Figures 8-10. Visual inspection reveals complex, nonlinear relationships between perturbations in many features and predicted ADRD risk. For lab and vital measurements, changes in serum anion gap, folate, mean corpuscular volume, and systolic blood pressure levels generally lead to increased risk, with increments showing a slightly greater impact than decrements. Conversely, changes in body mass index, body surface area, body weight, glomerular filtration rate, and triglycerides tend to reduce predicted risk, with increments showing slightly stronger protective effects. Clear positive correlation (i.e. increment leads to higher risk, decrement leads to lower risk) were observed for serum basophils volume, calcium, cholesterol (mass), and neutrophils per 100 leukocytes, while clear negative correlation (i.e. increment leads to lower risk, decrement leads to higher risk) were seen for hematocrit volume, hemoglobin (mass), lymphocytes volume, and neutrophils (absolute count). Among CCS codes, the strongest risk-increasing diagnoses included Parkinson’s disease and conditions related to delirium, dementia, and other cognitive disorders. For risk medications, antipsychotics, antiparkinsonian agents, and antidepressants were the top three categories most strongly associated with changes in ADRD risk.

### Model interpretability at individual level

In this section, we present two ADRD patients from the held-out dataset as illustrative examples to demonstrate how GRU-D-RETAIN interprets the contributions of input features over time during follow-up. The predicted ADRD risks for these two patients are shown in Supplementary Figures 11 and 12. Patients A and B received ADRD diagnoses in the 7.5th and 9.5th year of follow-up, with notable increases in predicted risk occurring at 4 and 7 years, respectively. The figures also highlight the consistent prediction generated by both the GRU-D-RETAIN and GRU-D models across all six cross-validation folds.

Figure 7 presents GRU-D-RETAIN interpretations for Patient A at 4.5 and 8 years of follow-up. Over time, the model’s timestep-level attention evolved from focusing solely on the 4.5-year mark to a broader distribution across 4.5, 6, and 8 years. At the modality level, the early rise in predicted risk at 4.5 years was primarily driven by demographic information, CCS codes, and lab/vital measurements. At the feature level, key contributors included female gender (DEMO: gender_1), non-Hispanic ethnicity (DEMO: ethn_1), age above 60s, diagnoses such as hyperlipidemia, spondylosis, mood disorders, and other female genital disorders, along with functional impairment in food preparation (FSA: foodpreparing). Most of the top CCS-related features identified at 4.5 years continued to rank highly at 8 years, with their influence generally increasing over time.

Supplementary Figure 13 displays the GRU-D-RETAIN interpretations for Patient B. The top contributing features at 7.5 years of follow-up include demographic variables and diagnoses such as delirium, dementia, and other cognitive disorders, as well as essential hypertension and hyperlipidemia. Unlike Patient A, whose timestep-level attention shifted toward the 8th year near the endpoint, Patient B’s attention remained primarily focused on the 7.5-year mark, which carried the highest attention weight.

### Interpretations across training folds

Figure 8 illustrates how feature contribution interpretations can vary across different training folds for Patients A and B. For Patient A at 4 years of follow-up (Figure 8a), model folds 1, 3, and 5 produced generally consistent interpretations, particularly regarding contributing CCS codes. In contrast, folds 2, 4, and 6 yielded differing interpretations. By the 7.5th year (Figure 8b) —when Patient A received an ADRD diagnosis—folds 1, 3, 4, 5, and 6 consistently highlighted CCS codes such as delirium, dementia, and other cognitive disorders, along with essential hypertension, as key contributors besides demographic factors. However, fold 2 provided a markedly different interpretation, as it assigned most of the timestep-level attention to the 5.5-year mark instead.

**Figure 8:**
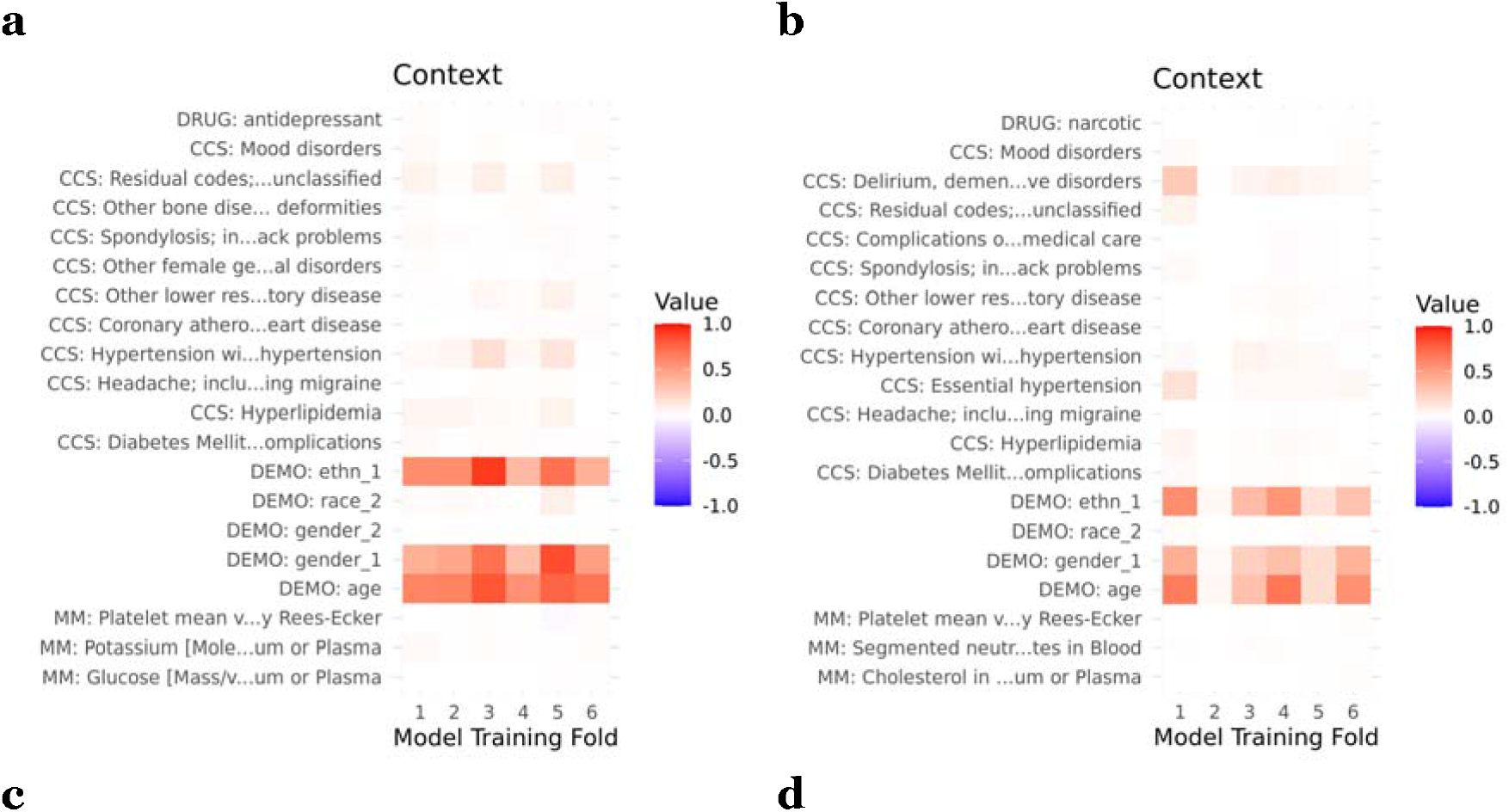
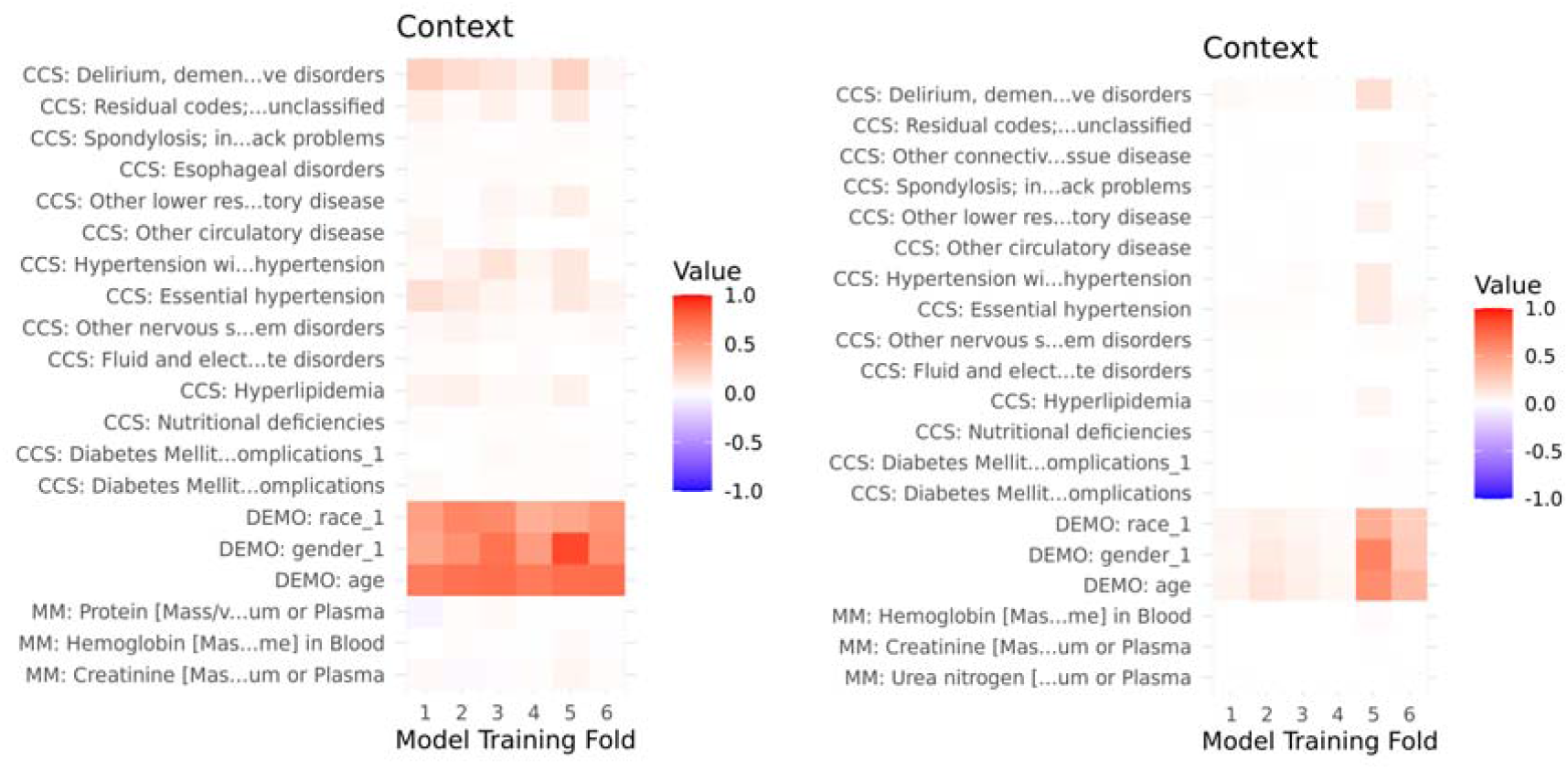
Context of the top 20 contributing features across the 6 training folds for a) patient A at 4 years, b) patient A at 7.5 years, c) patient B at 7 years, d) patient B at 9.5 years of follow-up.

For Patient B at the 7-year mark, all model folds produced generally consistent interpretations, with only minor variations in specific CCS codes. However, at 9.5 years, five out of six model folds continued to assign the highest timestep-level attention to the 7th year. As a result, only the fifth fold provided a visually distinct interpretation specific to the 9.5-year timepoint.

## Discussion

In this study, we proposed and evaluated GRU-D-RETAIN, a self-explainable longitudinal deep learning architecture for dynamic risk modeling of ADRD. The model was designed to address the challenges of sparse and irregular real-world EHR data by integrating the temporal modeling strength of GRU-D ^20^ with the interpretability of the RETAIN ^24^ attention mechanism. Despite the high level of data sparsity introduced through random time shifting—to reflect real-world clinical practice—GRU-D-RETAIN achieved predictive performance closely approximated the original GRU-D model over a 10-year follow-up period. Moreover, it offered clinically meaningful explanations of its predictions, with feature importance and dependence patterns largely consistent with established ADRD risk factors. These findings highlight the potential of GRU-D-RETAIN as a practical and interpretable solution for longitudinal risk monitoring in settings characterized by irregular clinical encounter and limited data availability, common challenges in population health and aging research.

A major challenge in EHR-based dynamic risk modeling for ADRD is the limited data availability during the period when prevention remains feasible—typically more than five years before diagnosis ^18,52^. Although cerebrospinal fluid (CSF)-derived biomarkers such as p-tau, t-tau, and Aβ42 have demonstrated predictive value as early as 20 years before diagnosis ^53^, none of these biomarkers were recorded in current ADRD cases and controls. Other CSF-related measurements, such as CSF glucose, CSF IgG, and CSF color, appear with extremely low presence rates—typically below 4 per 10,000 patients per year—which renders them unsuitable for training predictive models. Given these limitations, we propose that while EHR-based methods can establish ADRD risk profile up to 10 years prior to clinical diagnosis, additional clinical evaluation and specialized ADRD biomarker testing are necessary for narrowing down high-risk patients.

Our findings suggest that traditional logistic regression models, while reported effective in detecting existing ADRD cases ^54^, are inadequate for dynamic risk monitoring using EHR data. In our experiments, training logistic models on either dynamically sampled timepoint-specific data or cross-sectional data anchored at the index date resulted in poor predictive performance. This is primarily due to data sparsity across much of the pre-diagnosis period and the randomization of patients’ clinical encounters. We also emphasize that approaches which train and evaluate models using a fixed-length prediction window before clinical ADRD diagnosis ^12,14^, which assumes prior knowledge of the ADRD diagnosis date (a condition can not be met in real-world clinical settings), may lead to overly optimistic performance estimates and limit clinical applicability. Notably, even with a simpler imputation method, the LSTM model’s performance improves over time, easily outperforming the logistic models. This suggests that critical predictive information is embedded within the longitudinal progression of the ADRD, underscoring the necessity of employing temporal models for the ADRD risk prediction.

Permutation feature importance and PDP analysis suggest that the logic behind GRU-D-RETAIN model’s predictions are largely consistent with established knowledge of ADRD risk factors. Here, we highlight several observations that warrant further discussion. First, the unclassified residual code emerges as a key factor, second only to the diagnosis of cognitive disorders. This category encompasses over 300 diverse ICD-9 codes, including conditions such as sleep disorders, memory loss, and family history of disease. Further analysis would be valuable to identify which specific factors within this group contribute most significantly. Second, demographic features, represented as static variables replicated across all timesteps, consistently dominate the attention-based interpretations, often showing abrupt increases in contribution at specific timesteps when new CCS codes appear. A plausible explanation is that the demographic features are more stable inputs than temporally sparse CCS codes, and exploited by the model as statistical shortcuts during training. For example, if ADRD diagnoses frequently co-occur with older, non-Hispanic female patients, the model may primarily attribute increased risk to demographics rather than the diagnostic codes. Third, PDP revealed most of the risk medications, such as antipsychotic drugs, when tuned to either present or absent, increase the ADRD risk compared to original predictions. As the research in this space primarily covers the adverse outcome of using such medications ^55^ or among people already have dementia diagnosis ^56^, further research is helpful to validate the current observation and clarify the clinical context for safe initiation and withdrawal.

By examining interpretations across different model training folds, we observed that although the predicted ADRD risks remain relatively consistent, the specific contributing timesteps and features potentially vary. This likely stems from a combination of the complex, multifactorial nature of ADRD as well as the sparsity of the input feature space, which gives the model high flexibility to arrive at similar predictions through different explanatory pathways. These findings highlight a broader concern about the reliability of attention mechanisms as explanatory tools—a topic that has been the subject of ongoing debate ^57,58^. In the context of complex phenotypes like ADRD, we suggest that attention-based frameworks may be useful in narrowing the range of plausible interpretations, but the ultimate interpretive authority should rest with clinicians. It also underscores the value of training multiple model folds—akin to consulting a panel of clinical experts—to yield a more comprehensive set of potential interpretations.

## Conclusion

Our study indicates that EHR can serve as a valuable data source for ADRD risk monitoring up to 10 years prior to clinical diagnosis. The practical utility of such models is highly dependent on the availability and completeness of patient data, which can vary significantly across individuals. The GRU-D-RETAIN framework offers a distinct advantage by enabling real-time risk monitoring while providing interpretability at both the timestep and feature levels. This makes it a powerful tool for assisting healthcare providers in identifying high-risk patients and uncovering potential relevant risk factors—not only for ADRD, but also for other conditions where dynamic risk assessment and transparent model outputs are essential for clinical decision-making.

### Limitations

This study is based on EHR from a single hospital site, and is therefore subject to the limitations inherent in single-site studies. These include limited generalizability to broader populations, potential site-specific practice patterns, and institution-dependent data quality or coding behaviors that may not reflect those of other healthcare systems. For illustration purposes, this experiment was designed by masking data prior to the initiation of follow-up and predicting the risk of ADRD within 10 years from that point forward. As a result, the prediction horizon dynamically decreases from 10 to 0 years as patients progress along the follow-up timeline. In real-world applications, it is possible to estimate risk at any desired prediction horizon between 0 and 10 years using one of several strategies: (1) truncate or pad the input sequence to match the desired horizon without altering the current data sampling method, (2) revise the data sampling approach to incorporate information from before follow-up initiation, or (3) develop separate models tailored to each prediction horizon. These strategies can be applied individually or in combination. A dedicated benchmarking process is needed to determine the optimal approach and to assess how data availability and the length of follow-up affect model performance.

## Declarations

### Funding

This work was funded by National Human Genome Research Institute (3R01HG012748-02S2), Cancer Prevention and Research Institute of Texas Established Investigator Award (RR230020), and National Institute of Aging (R01AG072799-03). The funder played no role in study design, data collection, analysis and interpretation of data, or the writing of this manuscript.

### Author Contributions

XR: Model design, data analysis, manuscript writing.

SL: Data analysis, cross-validation.

RL: Expert opinion on time-series model design.

SF, JA, FC: NLP-related data curation.

VT, EO: Expert opinion from a clinician’s perspective, manuscript revision.

AW: Data scientist supporting OMOP infrastructure.

LW,HL: Expert opinion on cohort selection and data analysis, project coordination, manuscript revision.

### Data Availability

The data used in this study contain sensitive patient information and, therefore, are not publicly available. Access to the data is restricted to protect patient privacy and confidentiality. Researchers interested in accessing the data for academic purposes may contact the corresponding author for more information on the terms and conditions for data access.

### Code Availability

The code for GRU-D model is publicly available at https://github.com/PeterChe1990/GRU-D

The code for GRU-D-RETAIN is publicly available at https://github.com/ruanxiaoyang-UT/GRU-D-RETAIN/tree/main

### Competing Interests

The authors declare no competing interests.

## Acknowledgements

Not applicable

## Notes

### Competing Interest Statement

The authors have declared no competing interest.

### Author Declarations

Institutional Review Board (IRB) of University of Texas gave ethical approval for this work (IRB Number: HSC-SBMI-24-0403).

## References

1. Li, X. et al. Global, regional, and national burden of Alzheimer’s disease and other dementias, 1990–2019. Front. Aging Neurosci. 14, 937486 (2022).

2. 2024 Alzheimer’s disease facts and figures. Alzheimers Dement 20, 3708–3821 (2024).

3. Villemagne, V. L. et al. Amyloid β deposition, neurodegeneration, and cognitive decline in sporadic Alzheimer’s disease: a prospective cohort study. Lancet Neurol 12, 357–367 (2013).

4. Mitchell, S. L. CLINICAL PRACTICE. Advanced Dementia. N Engl J Med 372, 2533–2540 (2015).

5. Kumar, S. et al. Machine learning for modeling the progression of Alzheimer disease dementia using clinical data: a systematic literature review. JAMIA Open 4, ooab052 (2021).

6. Borchert, R. J. et al. Artificial intelligence for diagnostic and prognostic neuroimaging in dementia: A systematic review. Alzheimers Dement 19, 5885–5904 (2023).

7. Javeed, A. et al. Machine Learning for Dementia Prediction: A Systematic Review and Future Research Directions. J Med Syst 47, 17 (2023).

8. Kim, J., Jeong, M., Stiles, W. R. & Choi, H. S. Neuroimaging Modalities in Alzheimer’s Disease: Diagnosis and Clinical Features. Int J Mol Sci 23, (2022).

9. Akter, S., Liu, Z., Simoes, E. J. & Rao, P. Using Machine Learning and Electronic Health Record (EHR) Data for the Early Prediction of Alzheimer’s Disease and Related Dementias. medRxiv 2024.12.09.24318740 (2024) doi:10.1101/2024.12.09.24318740.

10. Wang, T., Qiu, R. G. & Yu, M. Predictive Modeling of the Progression of Alzheimer’s Disease with Recurrent Neural Networks. Sci Rep 8, 9161 (2018).

11. Wang, J. et al. Augmented Risk Prediction for the Onset of Alzheimer’s Disease from Electronic Health Records with Large Language Models. (2024).

12. Schliep, K. C. et al. Predicting the onset of Alzheimer’s disease and related dementia using Electronic Health Records: Findings from the Cache County Study on Memory in Aging (1995-2008). Res Sq (2024) doi:10.21203/rs.3.rs-4414498/v1.

13. Hu, X. et al. Self-Explainable Graph Neural Network for Alzheimer Disease and Related Dementias Risk Prediction: Algorithm Development and Validation Study. JMIR Aging 7, e54748 (2024).

14. Li, Q. et al. Early prediction of Alzheimer’s disease and related dementias using real-world electronic health records. Alzheimers Dement 19, 3506–3518 (2023).

15. Chen, Z. et al. Predicting the Risk of Alzheimer’s Disease and Related Dementia in Patients with Mild Cognitive Impairment Using a Semi-Competing Risk Approach. Informatics 10, 46 (2023).

16. Groenwold, R. H. H. Informative missingness in electronic health record systems: the curse of knowing. Diagn Progn Res 4, 8 (2020).

17. Nguyen, M., Sun, N., Alexander, D. C., Feng, J. & Thomas Yeo, B. T. Modeling Alzheimer’s disease progression using deep recurrent neural networks. https://ieeexplore.ieee.org/document/8423955.

18. Li, R. et al. Early Prediction of Alzheimers Disease Leveraging Symptom Occurrences from Longitudinal Electronic Health Records of US Military Veterans. (2023).

19. Post-Hoc Explanations Fail to Achieve their Purpose in Adversarial Contexts. https://dl.acm.org/doi/10.1145/3531146.3533153 doi:10.1145/3531146.3533153.

20. Che, Z., Purushotham, S., Cho, K., Sontag, D. & Liu, Y. Recurrent Neural Networks for Multivariate Time Series with Missing Values. Sci. Rep. 8, 6085 (2018).

21. Ruan, X., Wang, L., Thongprayoon, C., Cheungpasitporn, W. & Liu, H. GRU-D-Weibull: A novel real-time individualized endpoint prediction. Artif Intell Med 146, 102696 (2023).

22. Ruan, X. et al. Real-time risk prediction of colorectal surgery-related post-surgical complications using GRU-D model. J Biomed Inform 135, 104202 (2022).

23. Ruan, X. et al. Revolutionizing Postoperative Ileus Monitoring: Exploring GRU-D’s Real-Time Capabilities and Cross-Hospital Transferability. medRxiv (2024) doi:10.1101/2024.04.24.24306295.

24. RETAIN: An Interpretable Predictive Model for Healthcare using Reverse Time Attention Mechanism. ar5iv https://ar5iv.labs.arxiv.org/html/1608.05745.

25. Hripcsak, G. et al. Observational Health Data Sciences and Informatics (OHDSI): Opportunities for Observational Researchers. Stud Health Technol Inform 216, 574–578 (2015).

26. Vassilaki, M. et al. Characterizing Performance Gaps of a Code-Based Dementia Algorithm in a Population-Based Cohort of Cognitive Aging. J Alzheimers Dis 95, 931–940 (2023).

27. Wang, V. & Carpenter, W. R. Healthcare cost and utilization project (HCUP). Encyclopedia of Health Services Research (2015) doi:10.4135/9781412971942.n164.

28. Fu, S. et al. FedFSA: Hybrid and federated framework for functional status ascertainment across institutions. J Biomed Inform 152, 104623 (2024).

29. Fu, S. et al. A hybrid model to identify fall occurrence from electronic health records. Int J Med Inform 162, 104736 (2022).

30. Fu, S. et al. Quality assessment of functional status documentation in EHRs across different healthcare institutions. Front Digit Health 4, 958539 (2022).

31. Liu, S. et al. An open natural language processing (NLP) framework for EHR-based clinical research: a case demonstration using the National COVID Cohort Collaborative (N3C). J Am Med Inform Assoc 30, 2036–2040 (2023).

32. Reitz, C. Dyslipidemia and the risk of Alzheimer’s disease. Curr Atheroscler Rep 15, 307 (2013).

33. Mendez, M. F. Degenerative dementias: Alterations of emotions and mood disorders. Handb Clin Neurol 183, 261–281 (2021).

34. Åström, D. O. et al. High risk of developing dementia in Parkinson’s disease: a Swedish registry-based study. Scientific Reports 12, 1–7 (2022).

35. Jang, H.-D. et al. Relationship between dementia and ankylosing spondylitis: A nationwide, population-based, retrospective longitudinal cohort study. PLoS One 14, e0210335 (2019).

36. Ikram, M., Innes, K. & Sambamoorthi, U. Association of osteoarthritis and pain with Alzheimer’s Diseases and Related Dementias among older adults in the United States. Osteoarthritis Cartilage 27, 1470–1480 (2019).

37. Sierra, C. Hypertension and the Risk of Dementia. Front Cardiovasc Med 7, 5 (2020).

38. Tampi, R. R., Tampi, D. J., Balachandran, S. & Srinivasan, S. Antipsychotic use in dementia: a systematic review of benefits and risks from meta-analyses. Ther Adv Chronic Dis 7, 229–245 (2016).

39. Beghi, E. & Beghi, M. Epilepsy, antiepileptic drugs and dementia. Curr Opin Neurol 33, 191–197 (2020).

40. Bosco, F. et al. Antiseizure Medications in Alzheimer’s Disease from Preclinical to Clinical Evidence. Int J Mol Sci 24, (2023).

41. Gray, S. L. et al. Cumulative use of strong anticholinergics and incident dementia: a prospective cohort study. JAMA Intern Med 175, 401–407 (2015).

42. Costello, H., Roiser, J. P. & Howard, R. Antidepressant medications in dementia: evidence and potential mechanisms of treatment-resistance. Psychol Med 53, 654–667 (2023).

43. Coupland, C. A. C. et al. Anticholinergic Drug Exposure and the Risk of Dementia: A Nested Case-Control Study. JAMA Intern Med 179, 1084–1093 (2019).

44. Nho, K. et al. Association of Altered Liver Enzymes With Alzheimer Disease Diagnosis, Cognition, Neuroimaging Measures, and Cerebrospinal Fluid Biomarkers. JAMA Netw Open 2, e197978 (2019).

45. Lu, Y. et al. Liver integrity and the risk of Alzheimer’s disease and related dementias. Alzheimers Dement 20, 1913–1922 (2024).

46. Yi-Ming Li, Yen-Ching Chen, Jen-Han Chen, Jeng-Min Chiou, Ta-Fu Chen, Liang-Chuan Lai. Association between mean corpuscular volume and cognitive impairment in an 8-year cohort study in the community-dwelling elderly. https://alz-journals.onlinelibrary.wiley.com/doi/abs/10.1002/alz.039 (2020).

47. Qiang, Y.-X. et al. Associations of blood cell indices and anemia with risk of incident dementia: A prospective cohort study of 313,448 participants. Alzheimers Dement 19, 3965–3976 (2023).

48. Lucke, J. A. et al. Vital signs and impaired cognition in older emergency department patients: The APOP study. PLoS One 14, e0218596 (2019).

49. Ye, J. et al. Thyroid dysfunction and risk of different types of dementia: A systematic review and meta-analysis. Medicine (Baltimore) 103, e39394 (2024).

50. Al-Thani, N. A., Stewart, G. S. & Costello, D. A. The Role of the Urea Cycle in the Alzheimer’s Disease Brain. J Neurochem 169, e70033 (2025).

51. Chang, H.-C. et al. Interplay between orthopedic interventions and dementia: Dementia risk after total knee replacement. Alzheimers Dement 20, 5810–5812 (2024).

52. Guo, J., Gao, B., Huang, Y. & Song, S. Trajectory of multimorbidity before dementia: A 24-year follow-up study. Alzheimers Dement. (Amst.) 16, e12523 (2024).

53. Lespinasse, J., Dufouil, C. & Proust-Lima, C. Disease progression model anchored around clinical diagnosis in longitudinal cohorts: example of Alzheimer’s disease and related dementia. BMC Med Res Methodol 23, 199 (2023).

54. Lazarova, S., Grigorova, D., Petrova-Antonova, D. & For The Alzheimer’s Disease Neuroimaging Initiative. Detection of Alzheimer’s Disease Using Logistic Regression and Clock Drawing Errors. Brain Sci 13, (2023).

55. Rogowska, M. et al. Implications of Adverse Outcomes Associated with Antipsychotics in Older Patients with Dementia: A 2011-2022 Update. Drugs Aging 40, 21–32 (2023).

56. Van Leeuwen, E. et al. Withdrawal versus continuation of long-term antipsychotic drug use for behavioural and psychological symptoms in older people with dementia. Cochrane Database Syst Rev 3, CD007726 (2018).

57. Jain, S. & Wallace, B. C. Attention is not Explanation. (2019).

58. Wiegreffe, S. & Pinter, Y. Attention is not not Explanation. (2019).

