## Supplementary Figures for "A Self-Explainable Dynamic Risk Monitoring Framework for Predicting Alzheimer’s Disease and Related Dementias"

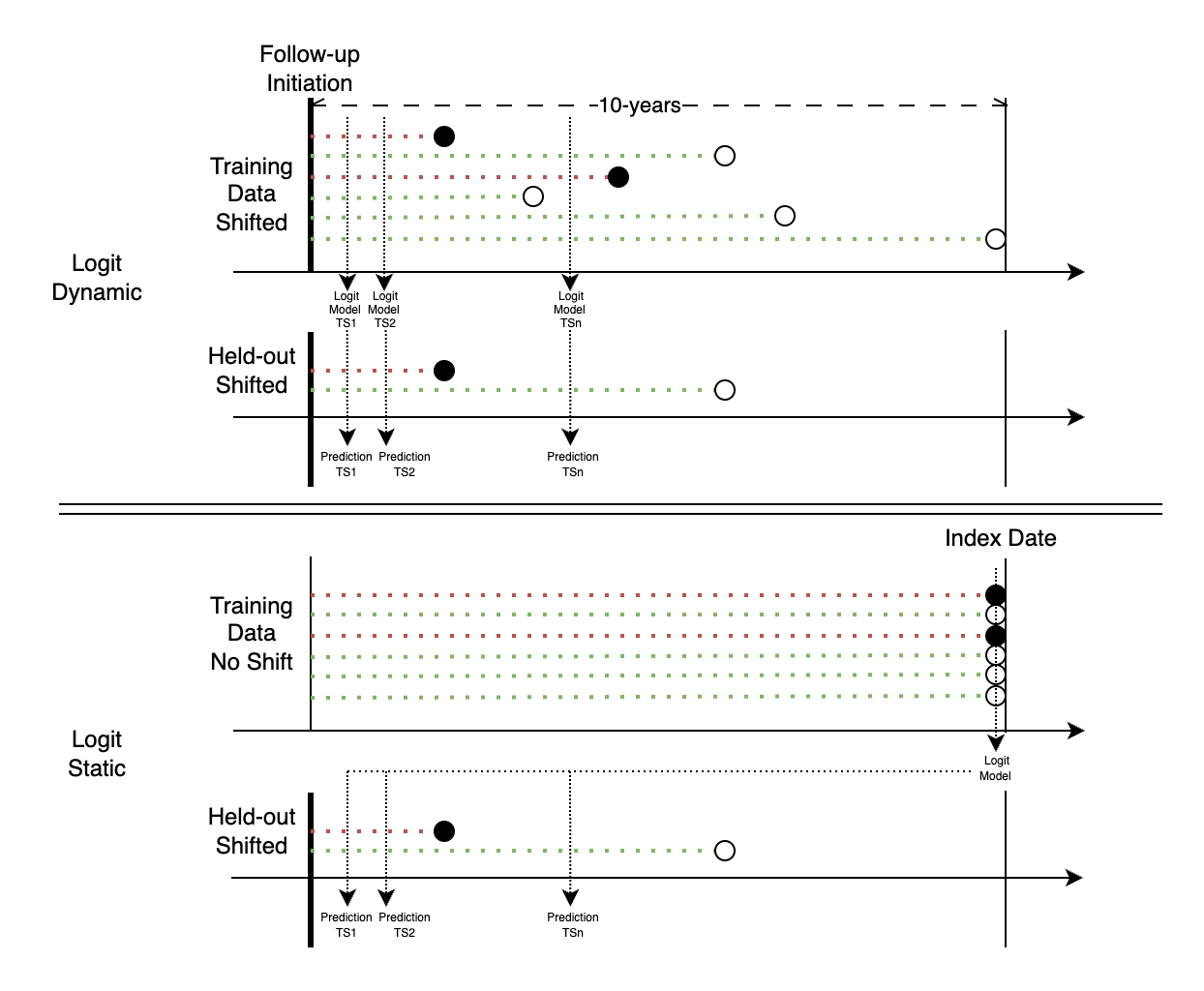

**Supplementary Figure 1** Illustration of Logit Dynamic and Logit Static models. Solid black circles represent ADRD cases. Hollow black circles represent controls.

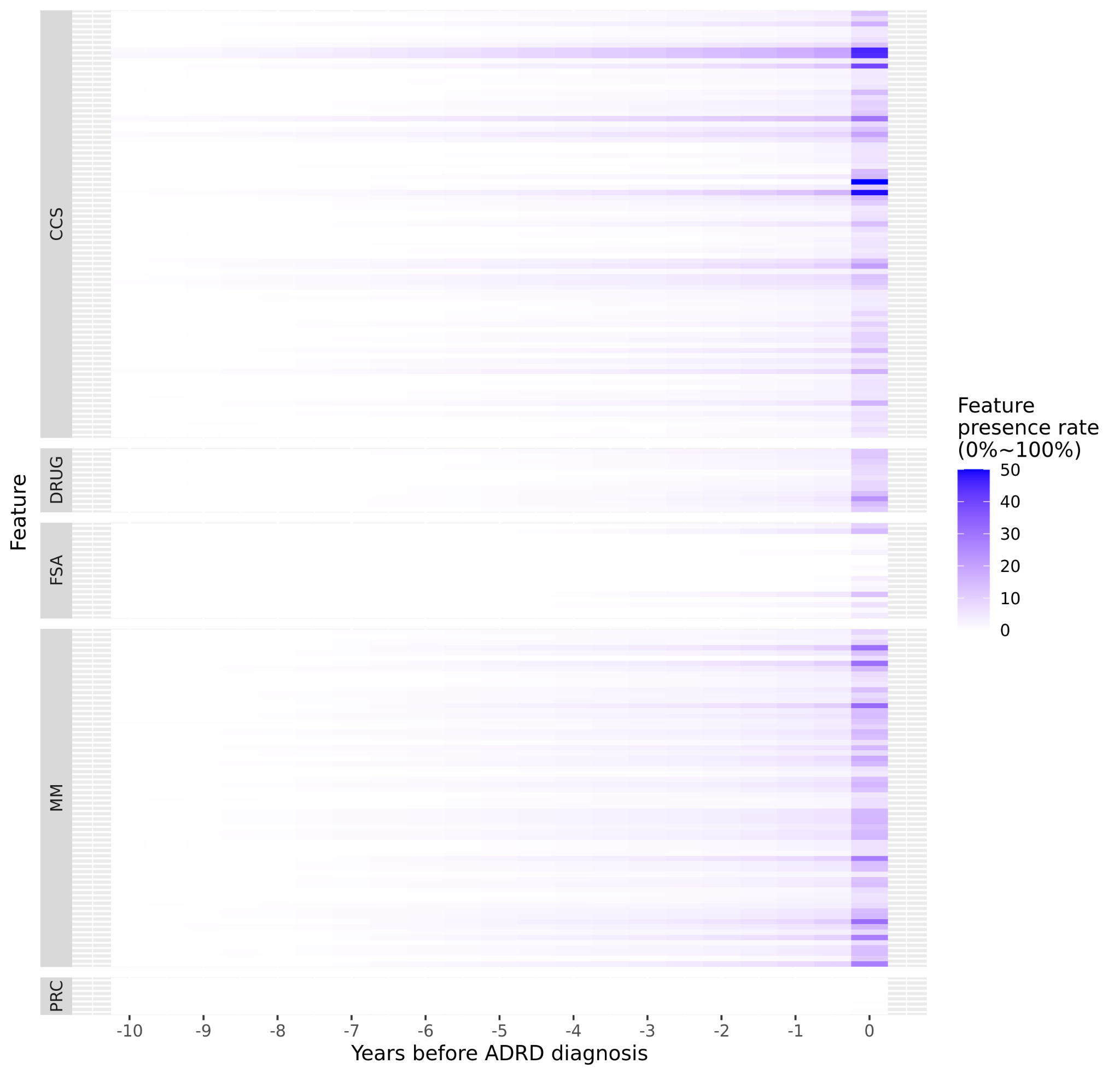

**Supplementary Figure 2** Data availability within 10 years before ADRD diagnosis. Each tile represents feature presence rate (0%~100%) among 14,937 ADRD patients within a half year period.

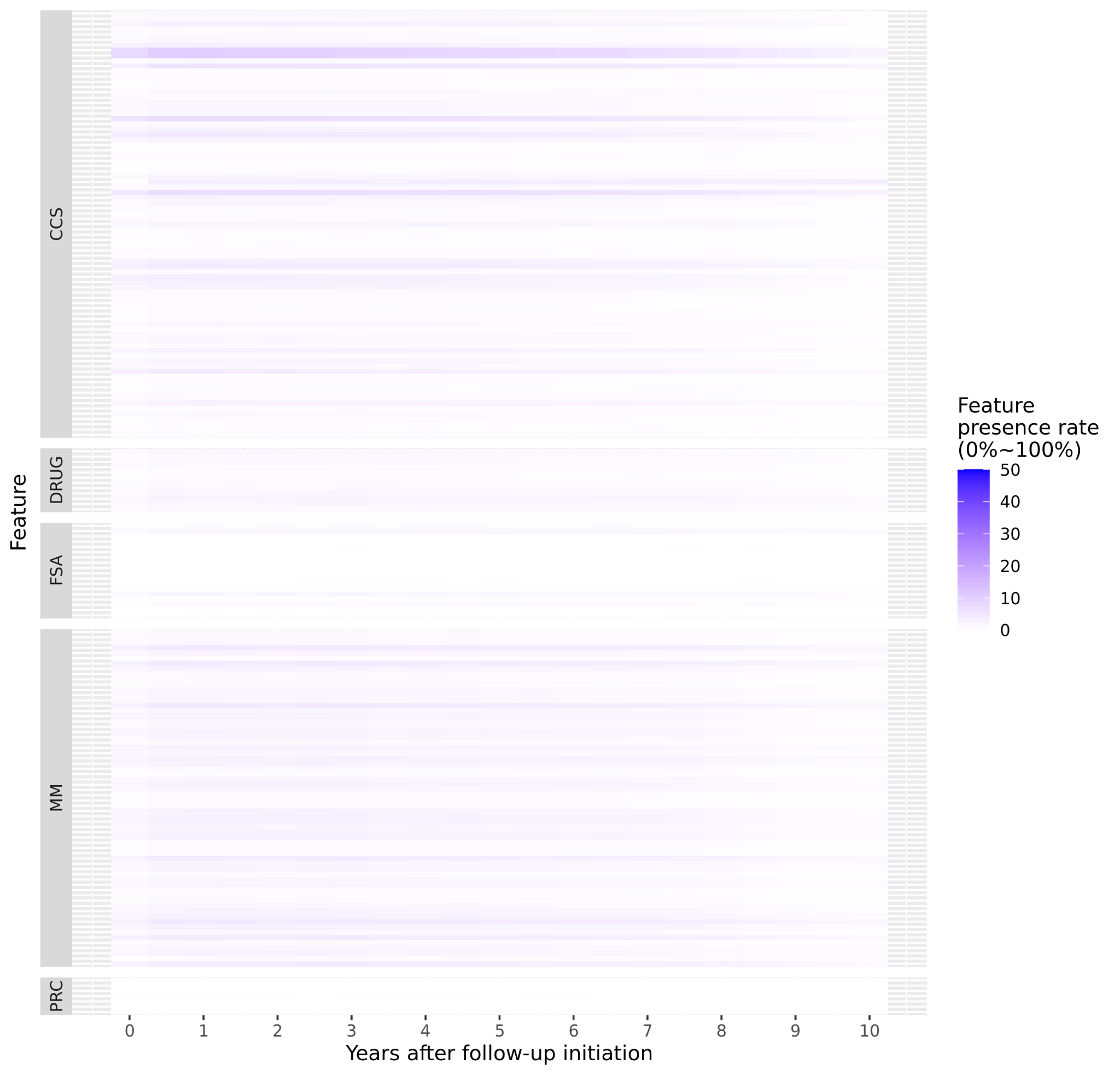

**Supplementary Figure 3** Data availability within 10 years after follow-up initiation. Each tile represents feature presence rate (0%~100%) among 14,937 ADRD patients within a half year period.

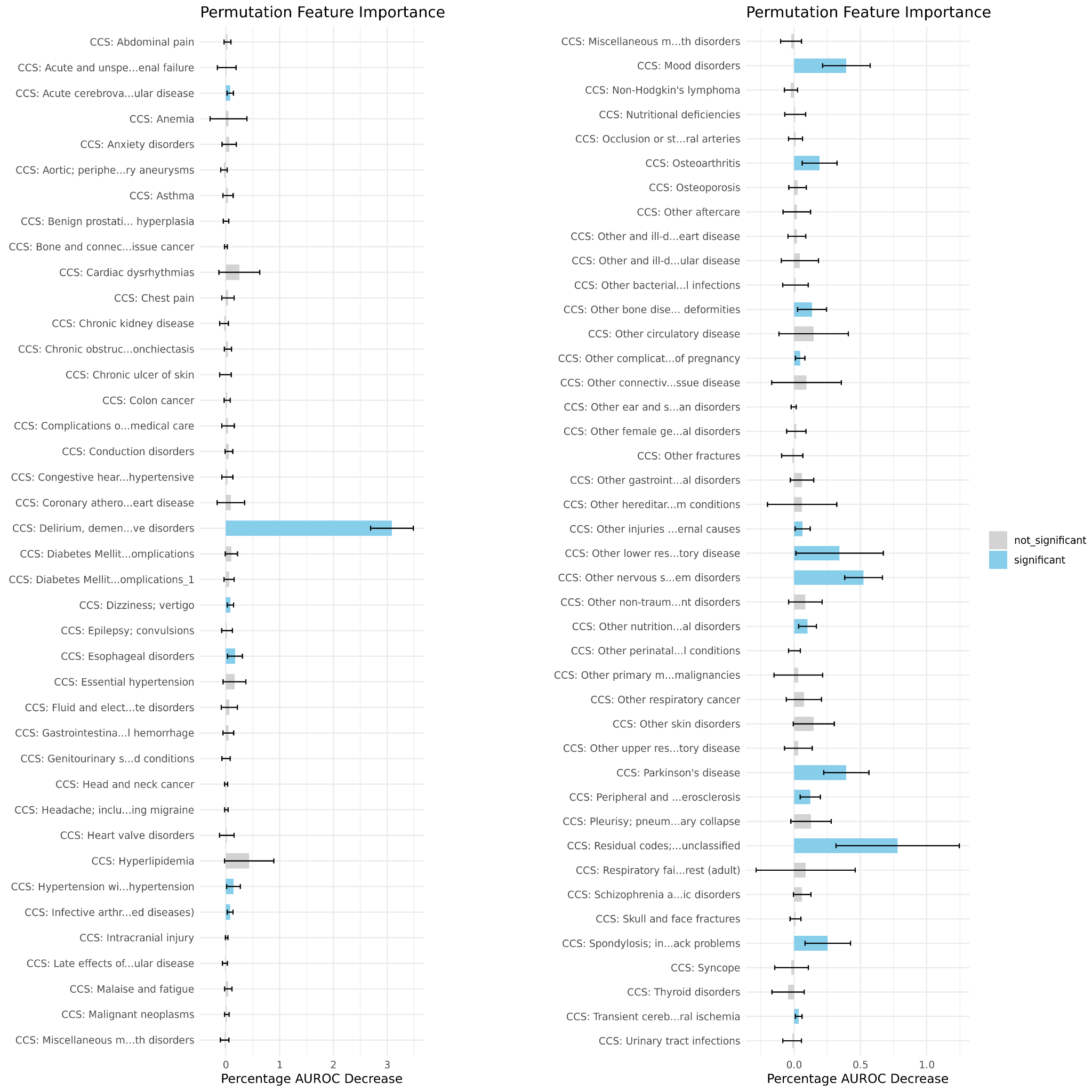

**Supplementary Figure 4** Permutation feature importance of 81 CCS codes consumed by GRU-D-RETAIN models. Error bars represent 95% CI based on 6-fold models.

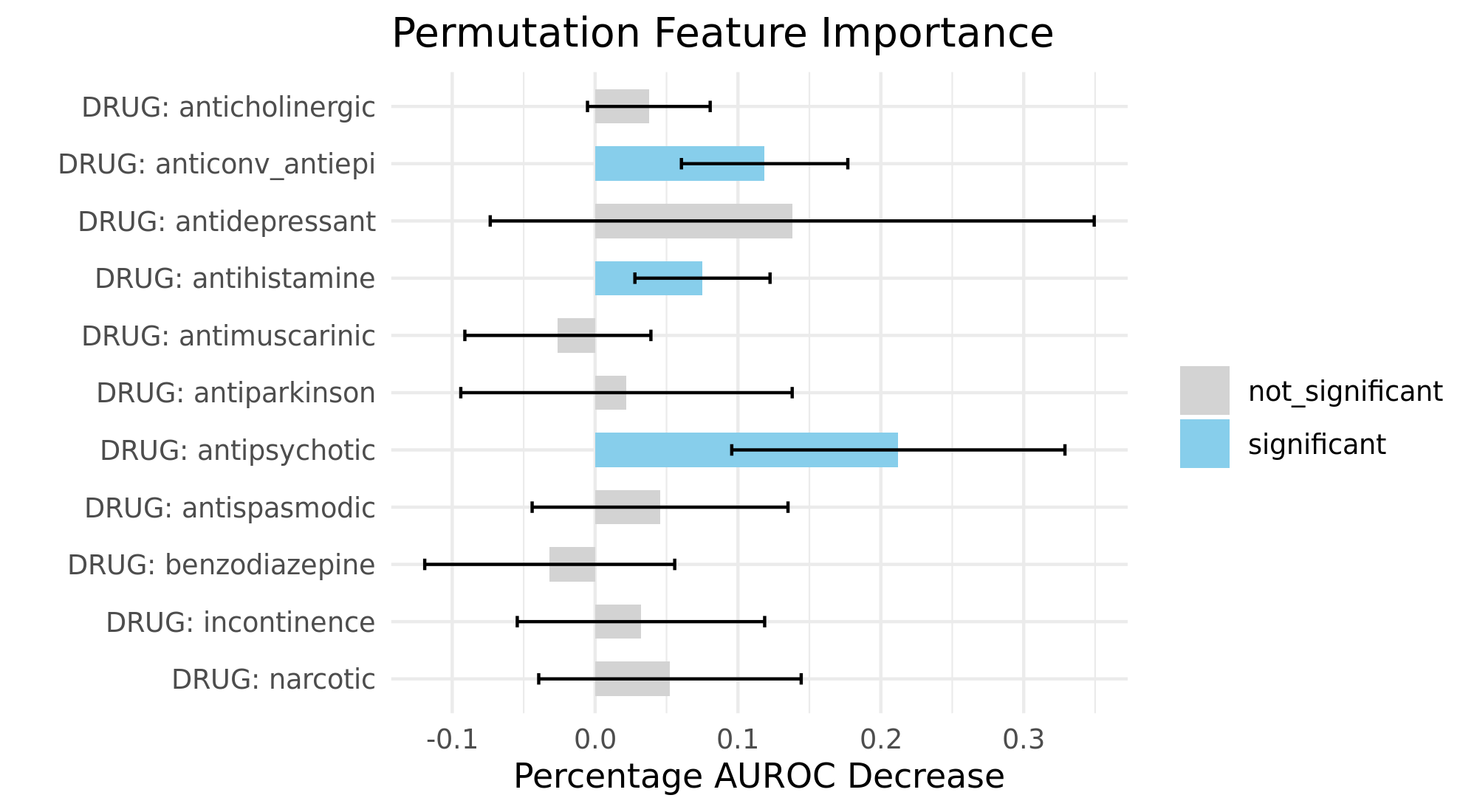

**Supplementary Figure 5** Permutation feature importance of 11 categories of potential risk medications consumed by GRU-D-RETAIN models. Error bars represent 95% CI based on 6-fold models.

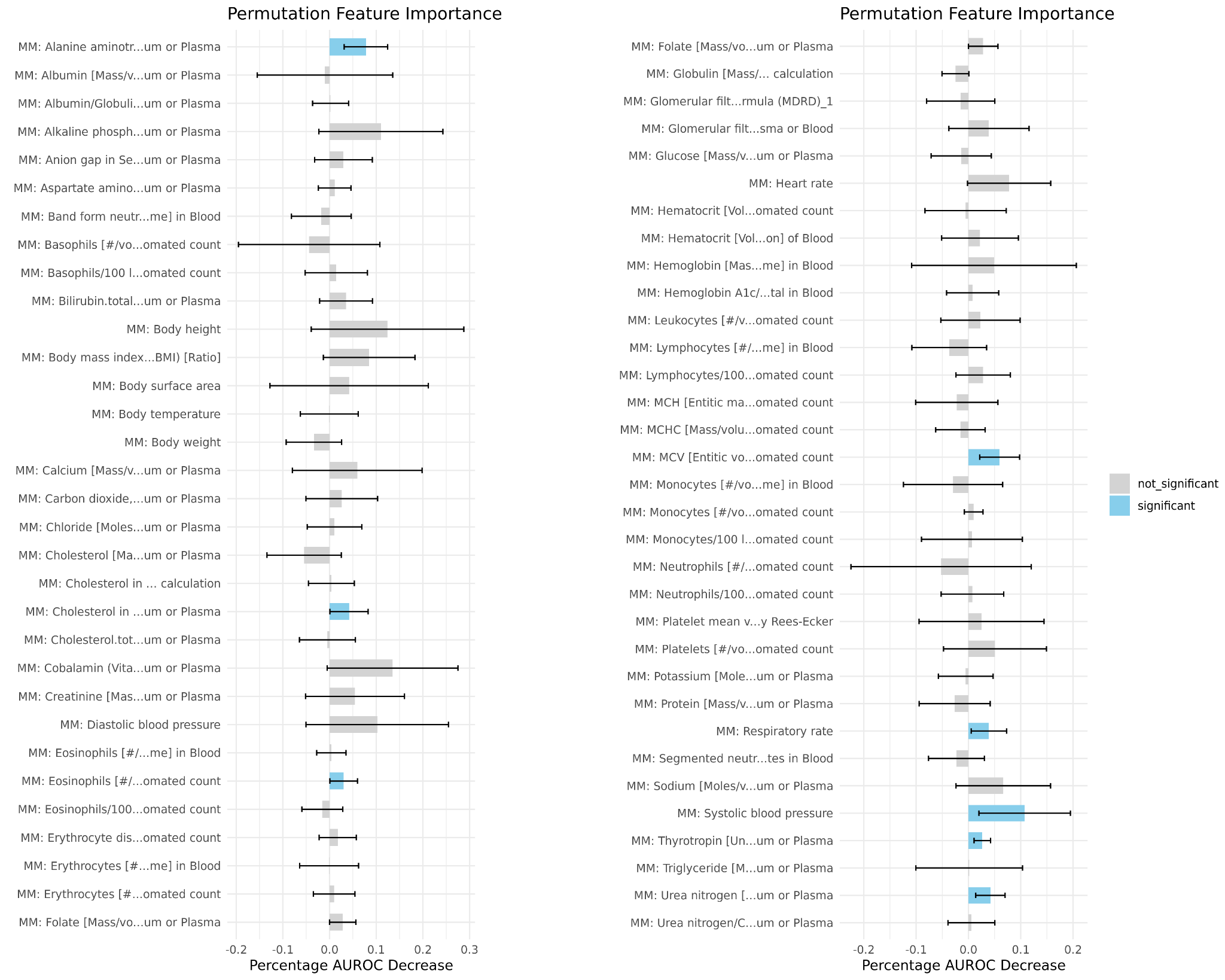

**Supplementary Figure 6** Permutation feature importance of 64 lab and vital measurements consumed by GRU-D-RETAIN models. Error bars represent 95% CI based on 6-fold models.

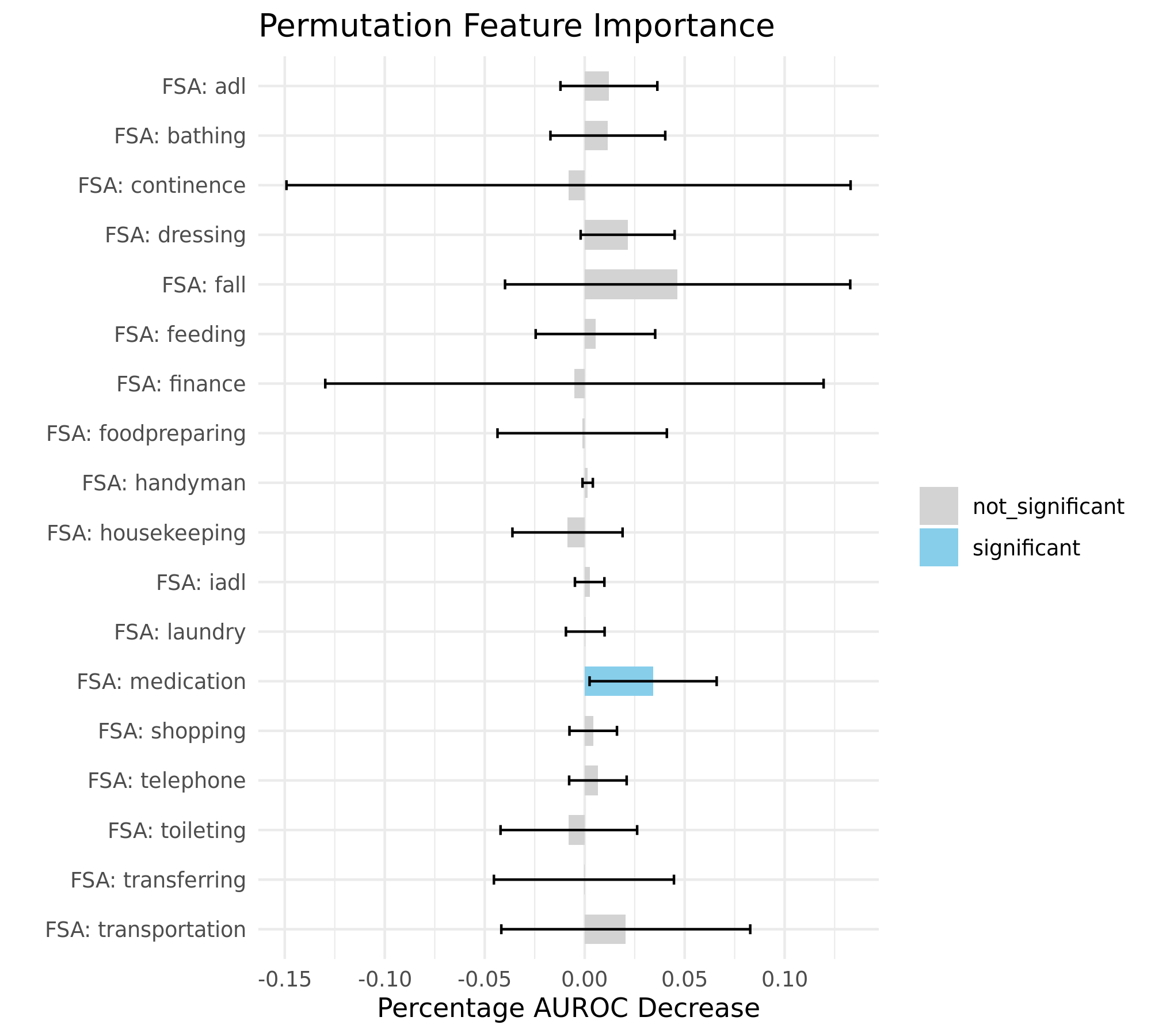

**Supplementary Figure 7** Permutation feature importance of FSA consumed by GRU-D-RETAIN models. Error bars represent 95% CI based on 6-fold models.

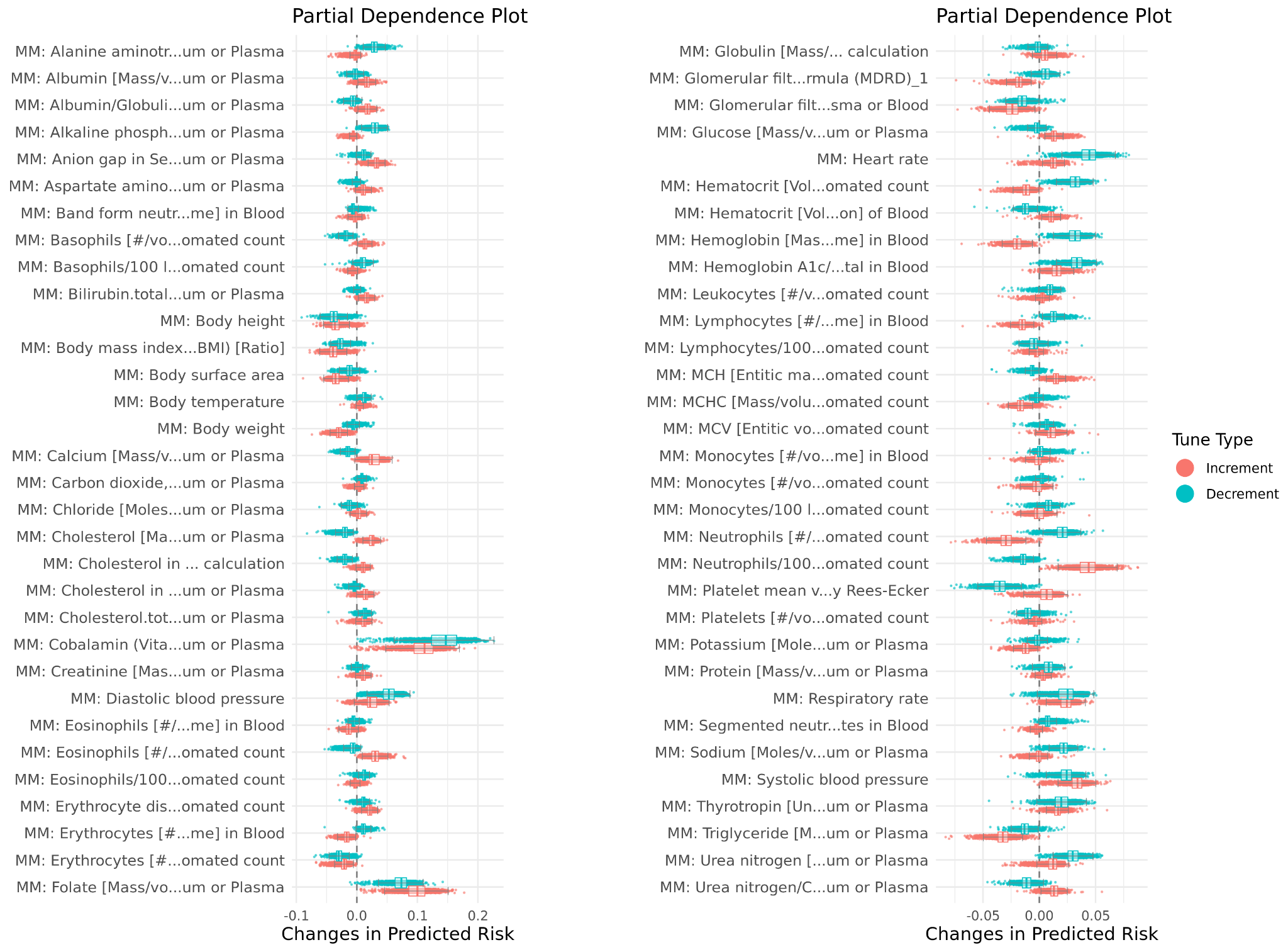

**Supplementary Figure 8** Partial dependence plots for 64 lab and vital measurements used by the GRU-D-RETAIN model. “Increment” and “decrement” indicate changes in predicted ADRD risk when each measurement is increased or decreased, respectively, by 1 standard deviation, compared to the original prediction. Results are based on 2,070 ADRD cases from the held-out dataset.

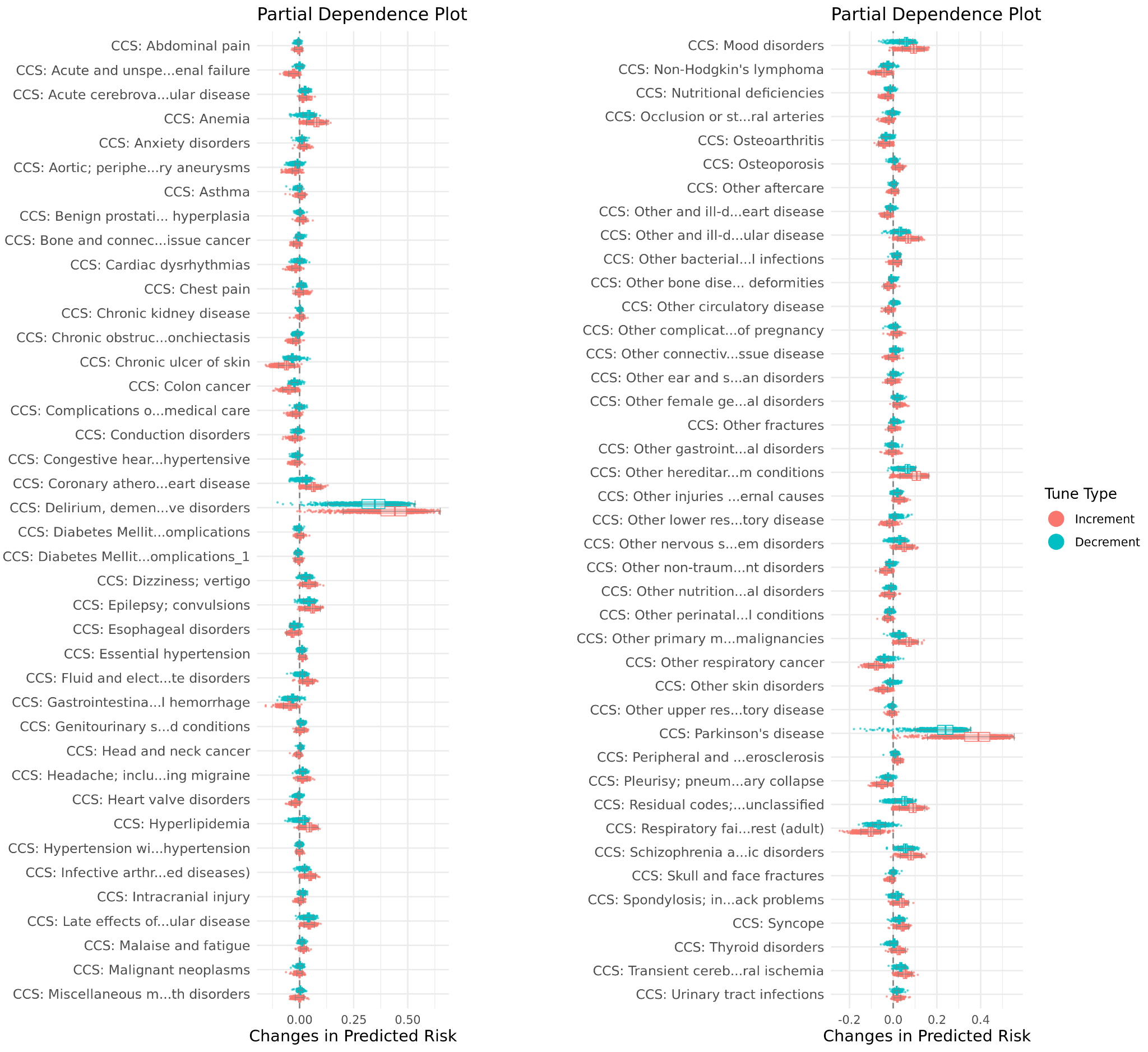

**Supplementary Figure 9** Partial dependence plots for 81 CCS codes used by the GRU-D-RETAIN model. “Increment” and “decrement” indicate changes in predicted ADRD risk when each CCS code is set to present or absent, respectively, compared to the original prediction. Results are based on 2,070 ADRD cases from the held-out dataset.

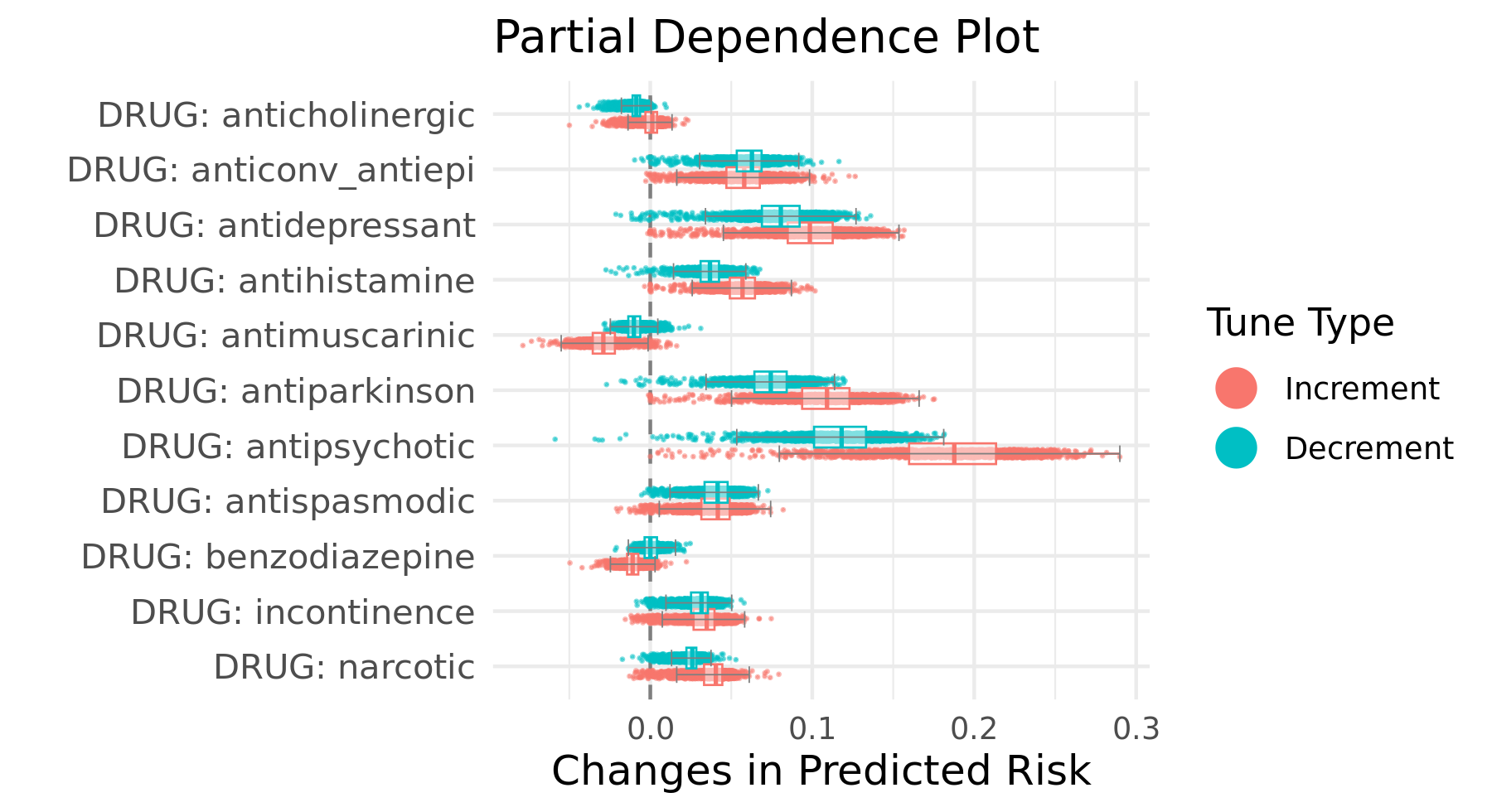

**Supplementary Figure 10** Partial dependence plots for risk medications used by the GRU-D-RETAIN model. “Increment” and “decrement” indicate changes in predicted ADRD risk when each medication is set to present or absent, respectively, compared to the original prediction. Results are based on 2,070 ADRD cases from the held-out dataset.

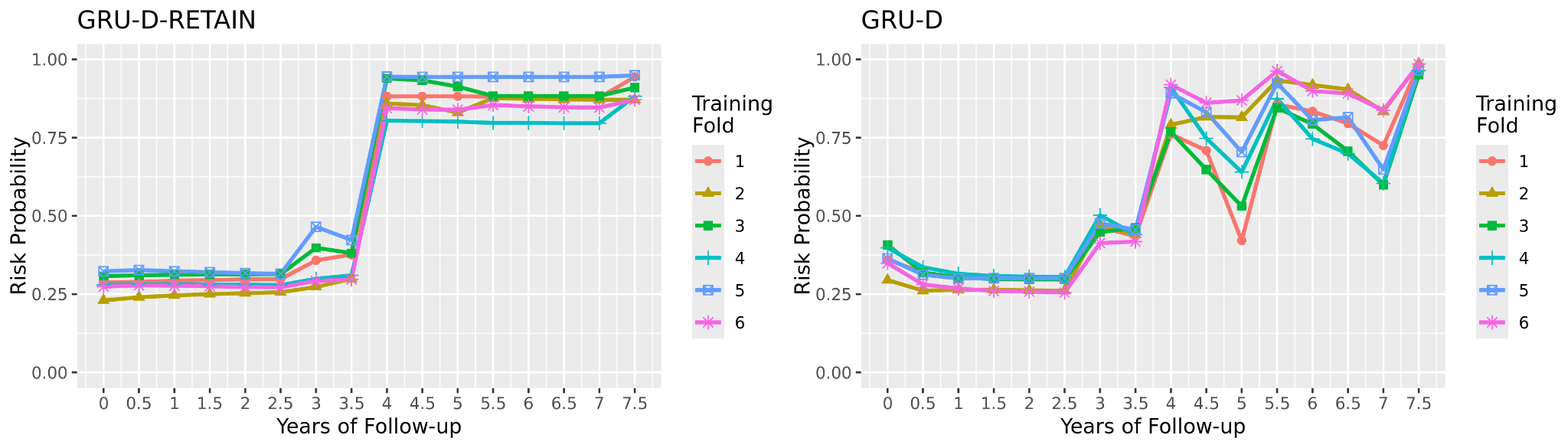

**Supplementary Figure 11** GRU-D-RETAIN and GRU-D based dynamic ADRD risk prediction for a patient with ADRD diagnosis at the 7.5th year of follow-up. Each color/shape represents prediction from one of the 6-fold models.

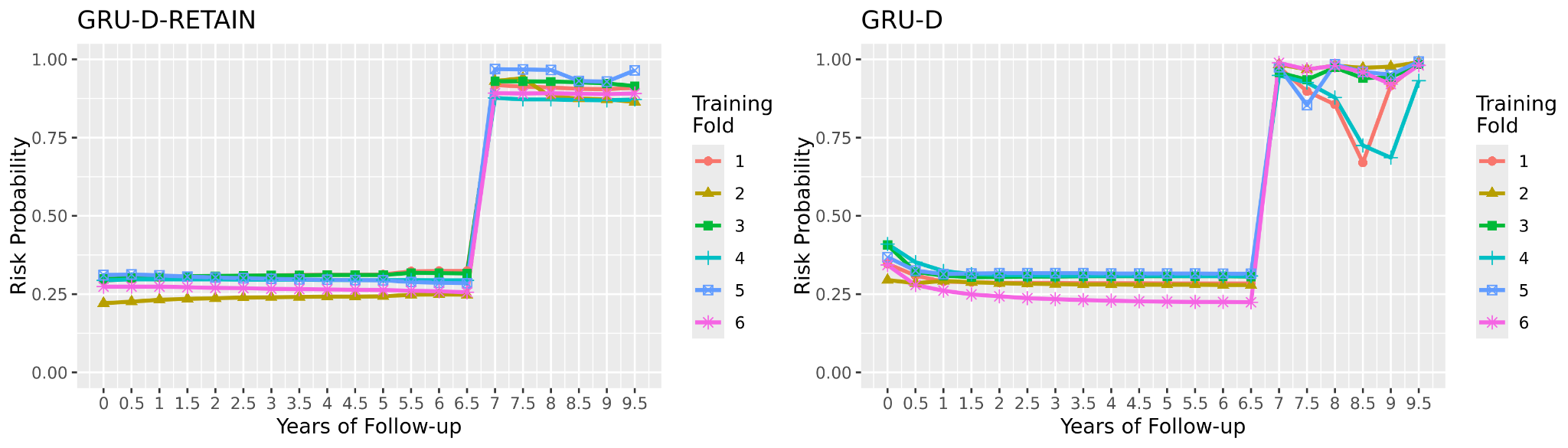

**Supplementary Figure 12** GRU-D-RETAIN and GRU-D based dynamic ADRD risk prediction for a patient with ADRD diagnosis at the 9.5th year of follow-up. Each color/shape represents prediction from one of the 6-fold models.

**a	b**

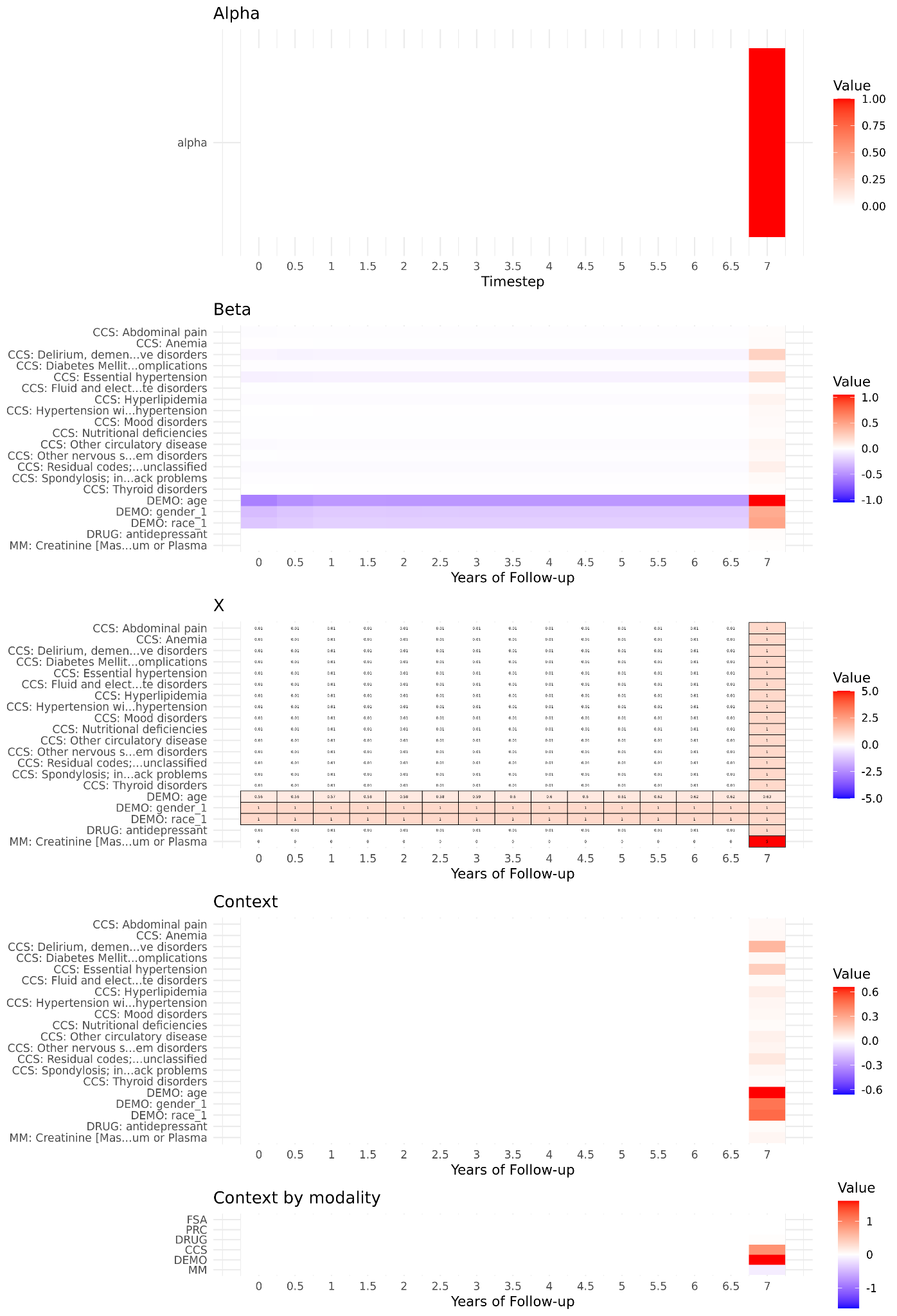

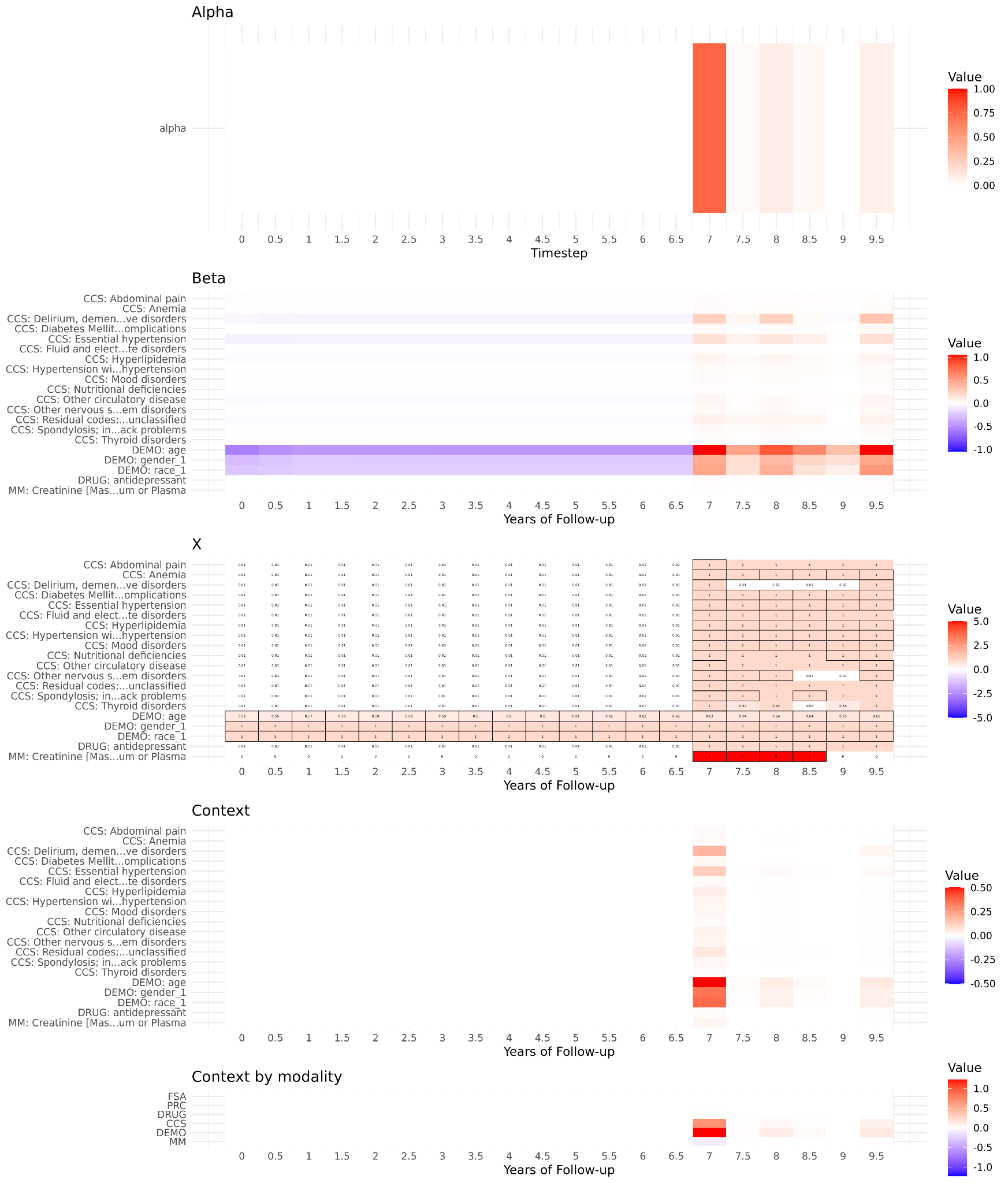

**Supplementary Figure 13** Interpretation for patient B with ADRD diagnosis at the 10th year of follow-up. a) Interpretation at 7.5 years, where the predicted risk significantly increases above the previous time interval. b) Interpretation at 10 years, where the patient had an ADRD diagnosis. Alpha: timestep level attention; Beta: feature level attention; X: values of input features; Context: Alpha*Beta*X. For input features (X), cells in black border indicate actual observed values, while others indicate missing observations imputed by GRU-D on the fly. Data shown for the top 20 features with largest contribution to the predicted risk.
